# Atypical energy-related symptoms define biologically distinct subtypes of major depressive disorder

**DOI:** 10.64898/2026.07.02.26357101

**Authors:** Arvid Harder, Rujia Wang, Jacob Bergstedt, Floris Huider, Siim Kurvits, Jackson Thorp, Tong Gong, Elham Assary, Anaïs B. Thijssen, Giuseppe Pierpaolo Merola, Anastasia A. Goula, Sam Bentwood, Robert Karlsson, Joelle A. Pasman, Chiara Fabbri, Ian B. Hickie, Wouter J. Peyrot, Douglas F. Levinson, Major Depressive Disorder Working Group of the Psychiatric Genomics Consortium, James B. Potash, Jianxin Shi, Sarah E. Medland, Brittany L. Mitchell, Alex S.F Kwong, Thalia C. Eley, Cathryn M. Lewis, Steven P. Hamilton, Nicholas G. Martin, Dorret I. Boomsma, Kelli Lehto, Naomi R. Wray, Gerome Breen, Brenda W.J.H. Penninx, Yuri Milaneschi, Sally Marshall, Pippa A. Thomson, Yi Lu

## Abstract

Major depressive disorder (MDD) is a complex psychiatric disorder, characterized by a range of mood, cognitive, and neurovegetative symptoms. Current diagnostic criteria treat opposite symptom directions as equivalent; weight gain or loss, and increased or decreased sleep, each count toward a single diagnosis. We defined three subgroups of individuals meeting criteria for MDD: *AERS+* (hypersomnia with increased appetite/weight gain), *AERS−* (insomnia with appetite/weight loss), and an Uncategorized group, and conducted genome-wide association meta-analyses for each (*N_eff_* = 47,858, 156,624, and 215,828, respectively). We identified 27 genome-wide significant loci across subtypes, 4 for *AERS+*, 10 for *AERS−* and 13 for Uncategorized. *AERS+* showed higher SNP-based heritability (10.9%) and lower polygenicity (1.7% of SNPs) than *AERS−* (7.9%; 2.9%) or Uncategorized (8.6%; 5.3%), with larger effect sizes at its associated loci. The *AERS+* and *AERS−* subtypes were moderately genetically correlated (*r*_g_= 0.64, se = 0.04). Metabolic traits emerged as a primary differentiator: *AERS+* correlated positively with BMI, metabolic syndrome, and related traits, whereas *AERS−* correlated weakly in the opposite direction. These findings show that the directionality of neurovegetative symptoms indexes genetic heterogeneity within MDD, with metabolic biology as a central axis of differentiation.

## Introduction

Major Depressive Disorder (MDD) remains a leading cause of global disease burden^1^, yet advances in treatment have stalled, in part due to the disorder’s profound heterogeneity. Individuals with the same MDD diagnosis manifest different symptom profiles that may represent the expression of partially distinct pathophysiological pathways^2^.

Diagnostic approaches to MDD have shifted from subtype-based formulations to a unitary diagnosis with optional specifiers in current classification. While this improved diagnostic standardization, it also institutionalized a research phenotype that aggregates individuals with opposing neurovegetative symptom directions - particularly appetite/weight change and sleep disturbance - despite longstanding clinical arguments that these patterns may index different underlying biology^3^. In the Diagnostic and Statistical Manual (DSM), version 3, melancholic (or “typical”) depression was conceptualized as a distinct subtype, characterized by insomnia, psychomotor retardation, and loss of appetite or weight, and was often regarded as the prototypical, biologically driven form of depressive illness. In contrast, “atypical” depression - defined by preserved mood reactivity, interpersonal rejection sensitivity, hypersomnia, increased appetite or weight gain, and leaden paralysis^4,5^ was recognized as a separate subtype with a preferential response to the monoamine oxidase inhibitor class of antidepressants^6^. In DSM-5-TR, these clinical distinctions were retained only as specifiers, reflecting persistent uncertainty regarding biological distinctions underlying these MDD subgroups.

Recent work has highlighted that MDD patients manifesting symptoms such as increased weight/appetite, hypersomnia and leaden paralysis - labelled “atypical energy-related symptoms (AERS)” - are characterized by chronic inflammation, abdominal obesity and metabolic dysfunction^7^. This has led to the hypothesis that these symptoms identify an immunometabolic subgroup of MDD^8^. However, genetic characterizations of these subtypes have been limited by small, underpowered studies^9–11^. Existing work has reported differences in BMI genetic liability between individuals with and without hypersomnia and weight gain^9^, but with conflicting findings on heritability differences across weight- and sleep-stratified subtypes^9,10^ and a single genome-wide significant locus distinguishing MDD cases with weight gain from controls^11^.

To remedy this, we used data from large population-based and clinical cohorts of European ancestry to perform the largest genetic analysis of AERS−defined MDD subgroups to date. We defined three subgroups based on direction of sleep and weight disturbances: *AERS+*, *AERS−*, and an Uncategorized group. By comparing the genetic architecture underlying these subgroups, our aim was to clarify a long-standing question in MDD nosology: whether heterogeneous neurovegetative profiles reflect different presentations of a single disorder or partially distinct entities masked by syndromic aggregation.

## Results

We harmonized diagnostic questionnaire data across eight sources of European ancestry samples in which directionality for weight/appetite and sleep symptoms during the worst lifetime major depressive episode was available (*Table 1*). We classified lifetime MDD by the worst lifetime episode if ≥5 of 9 DSM-5 symptoms were experienced for most of the day, nearly every day and were accompanied by moderate-to-severe functional impairment. We further subgrouped all MDD cases by energy-related symptoms during the worst episode. Cases with hypersomnia combined with increased appetite or weight gain were labelled AERS positive (*AERS+*), whereas those with insomnia/hyposomnia plus appetite or weight loss were labelled AERS negative (*AERS−)*. Individuals meeting criteria for MDD, but which did not fulfil the criteria for either *AERS*+ or *AERS−* were classified as Uncategorized. Uncategorized was the largest group, and *AERS−* was more prevalent than *AERS+* in all cohorts except AGDS (*Table 1*, *Figure 1A-B)*. The three subtypes differed in baseline BMI measures; *AERS+* had higher BMI compared to both subtypes as well as controls, while *AERS−* had lower BMI than other subtypes and controls (*Supplementary Figure 1*). To assess whether the AERS subtypes simply represent MDD cases stratified by BMI, we leveraged UK Biobank data and compared *AERS+* to BMI- and sex-matched MDD cases. Compared to the matched cases, *AERS+* was associated with earlier onset, higher recurrence, greater functional impairment, and elevated psychiatric and somatic comorbidity, indicating that this subtype is not explained by a different BMI profile alone (*Supplementary Note 1*, *Supplementary Figures 2–3 & Supplementary Table 1*).

**Figure 1;.**
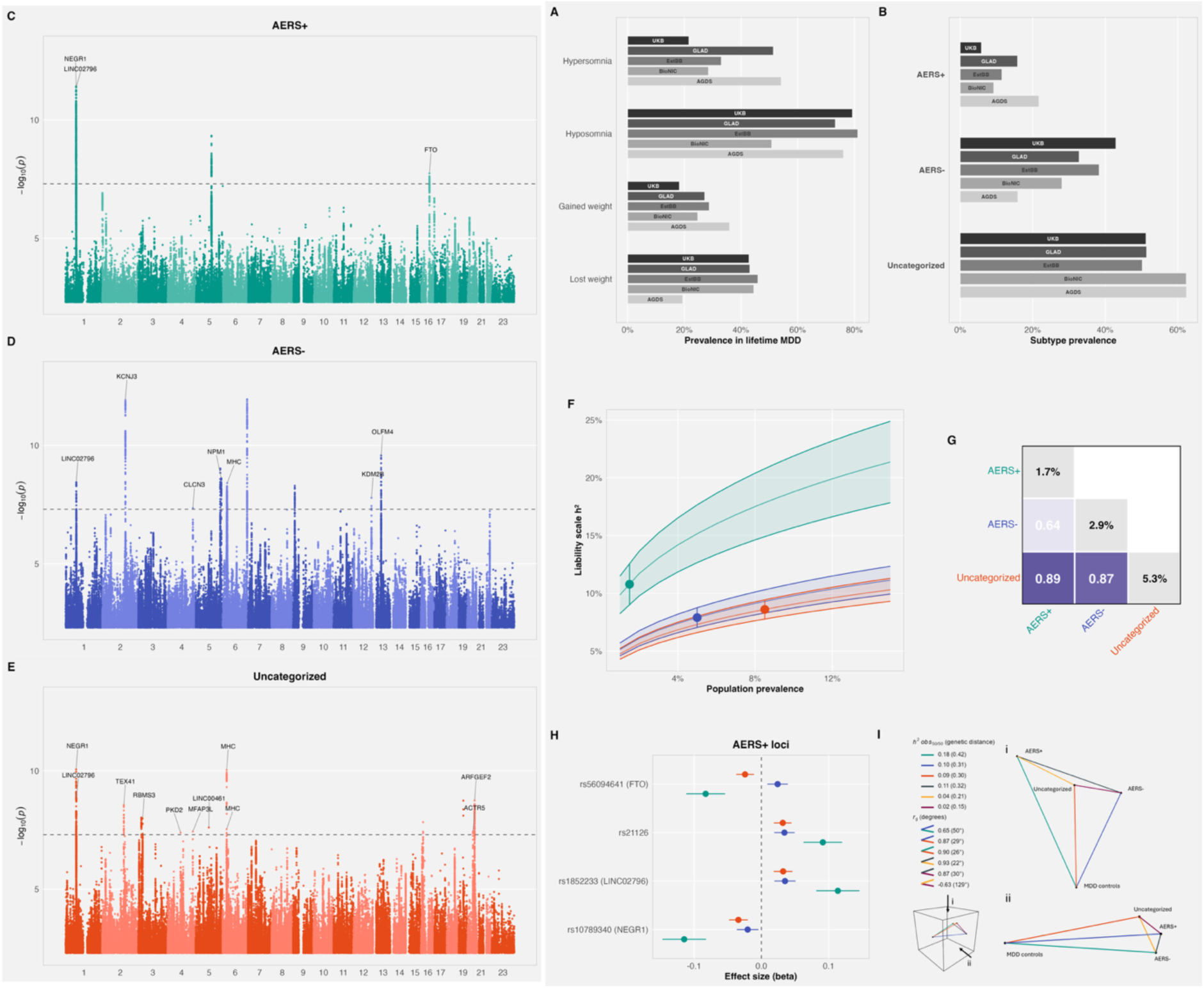
**(A)** Prevalence of weight and sleep symptoms (lost weight, gained weight, hyposomnia, hypersomnia) among lifetime MDD cases in the five largest contributing cohorts (AGDS, BioNIC, EstBB, GLAD, UKB). Bars are grouped by symptom; cohort identity is shown by greyscale shade and by the in-bar cohort label. **(B)** Subtype prevalence within lifetime MDD by cohort. For each cohort, the fraction of MDD cases classified as AERS+, AERS−, or Uncategorized is shown. **(C–E)** Manhattan plots of genome-wide association results for (C) AERS+, (D) AERS−, and (E) Uncategorized MDD. Each point is a SNP, plotted as −log₁₀(P) against genomic position. The dashed line marks the genome-wide significance threshold (P = 5 × 10⁻⁸). Lead variants at genome-wide significant loci are annotated by a nearby gene; the MHC region on chromosome 6 is collapsed to a single “MHC” label. **(F)** SNP-based heritability on the liability scale as a function of assumed population prevalence, for each subtype. Shaded ribbons indicate 95% confidence intervals; point ranges mark the estimate at the prevalence used in downstream analyses. **(G)** Pairwise genetic correlations (*r*_g_) between MDD subtypes (lower triangle) with SBayesS polygenicity estimates (Pi, percentage of SNPs with non-zero effects) on the diagonal. Tiles are colored by *r*_g_ magnitude. **(H)** SNP effect sizes (β) at AERS+ lead loci (rs10789340, rs1852233, rs21126, rs56094641) across all three subtypes. Error bars are 95% confidence intervals; the dashed line marks β = 0. **(I)** GDIS geometric representation of AERS+, AERS−, Uncategorized and controls. Subtype-subtype heritabilities and genetic correlations are based on GDIS transformations. Each line represents a GWAS; line lengths are based on genetic distance; angles between lines represent the genetic correlation between the GWASs; i = top view of 3D geometrical shape; ii = side view

**Table 1:** The table lists the cohorts analyzed, country of origin, and number of cases and controls for each AERS subtype. Relative prevalence of AERS+, AERS− and uncategorized is shown in parenthesis for each country.

| Cohort | Country | N_MD<br>D | N_AERS_<br>PLUS | N_AERS_<br>MINUS | N_UNCA<br>T | N_Contr<br>ols |
| --- | --- | --- | --- | --- | --- | --- |
| Genetic links to Depression and Anxiety and UK Biobank (GLAD UKBB) | UK | 68,494 | 5,832 (8.5%) | 27,649 (40.4%) | 35,013 (51.1%) | 225397 |
| The Biobanks Netherlands Internet Collaboration (BIONIC) | Netherlands | 17,092 | 1,583 (9.3%) | 4,812 (28.2%) | 10,697 (62.6%) | 48286 |
| Estonia Biobank (Estbb) | Estonia | 14,981 | 1,740 (11.6%) | 5,706 (38.1%) | 7,535 (50.3%) | 31391 |
| The Australian Genetics of Depression Study (AGDS) | Australia | 12,718 | 2,895 (22.8%) | 1,972 (15.5%) | 7,851 (61.7%) | 13138 |
| Psychiatric Genomics Consortium (PGC) | U.K, U.S | 5573 | 338 (6.1%) | 2,300 (41.3%) | 2,935 (52.7%) | 4644 |
| Swedish Twin Registry (STR) | Sweden | 2496 | 147 (5.9%) | 1,357 (54.4%) | 992 (39.7%) | 6817 |
| Twins early development study (TEDS) | U.K | 1760 | 212 (12.0%) | 634 (36.0%) | 914 (51.9%) | 3079 |
| Generation Scotland | U.K | 1234 | 105 (8.5%) | 480 (38.9%) | 649 (52.6%) | 3862 |
| **Total** |  | 124,348 | 12,852 (10.3%) | 44,910 (36.1%) | 66,586 (53.5%) | 336614 |

We conducted three meta-analyses of genome-wide association studies (GWAS), comparing *AERS+* (*N_eff_* = 47,858, N cases = 12,852, N controls = 334,919), *AERS−* (*N_eff_* = 156,624, N cases = 44,910, N controls = 336,614) and *Uncategorized* (*N_eff_* = 215,828, N cases = 66,586, N controls = 336,614), using a largely overlapping set of controls. LD score regression (LDSC) estimates of intercept indicated little to no residual population confounding (range: 0.990 to 1.019, *Supplementary Table 2-3*). We detected 27 (23 unique) genome-wide significant loci across the three subtypes: four for *AERS+*, 10 for *AERS−* and 13 for Uncategorized (*Figure 1C-E, Supplementary Table 4*). Most, but not all, of these genome-wide significant loci have been reported previously, either in the most recent MDD GWAS^12^ or in GWAS of other depression-related phenotypes (*Supplementary Table 4*).

### Atypical energy-related subtypes with partial overlap in genetic architecture

We observed higher estimates of SNP-based heritability (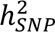 on the liability scale) across a range of population prevalences for *AERS+* compared to *AERS−* and Uncategorized (*Methods, Figure 1F*). Assuming population prevalences of 1.5%, 5.0% and 8.5% for *AERS*+, *AERS−*, and Uncategorized (*Supplementary Figure 4*), 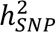 for *AERS+* was 10.9% (se = 0.91%), *AERS−*: 7.9% (se = 0.4%), and Uncategorized: 8.6% (se = 0.42%) (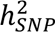 on the observed 50-50 scale^13^ were: 18%, 9.3%, 8.6% for *AERS*+, *AERS−*, and *Uncategorized*, respectively). We estimated pairwise genetic correlations (*r*_g_) using LDSC between the three AERS subtypes and observed a significantly lower genetic correlation between *AERS+* and *AERS−* (*r*_g_= 0.64, se = 0.04, *Figure 1G*) compared to their *r*_g_with *Uncategorized (e.g., AERS+ and AERS− compared to Uncategorized:* Δ*r*_g_= −0.25, se = 0.041, p =1.9 × 10^−9^). Polygenicity estimates using SBayesS (all estimates converged; Gelman-Rubin statistic < 1.2) indicated a smaller set of causal variants for *AERS+* (1.7% of all SNPs, posterior standard error 0.72%), compared to *AERS−* (2.92%, posterior standard error 0.59%) and *Uncategorized* (5.34%, posterior standard error 1.02%, *Figure 1G*). Consistent with higher heritability distributed across fewer causal variants, we observed stronger effect sizes at all four genome-wide significant loci for *AERS+* (*Figure 1H*). Transformation of the subtypes to geometrical space using GDIS^13^ revealed that the genetic distance from *AERS+* to *AERS−* was larger than the genetic distance from *AERS−* to controls and that the Uncategorized subtype is partially, but not fully, a genetic mix of *AERS+* and *AERS−* (*Figure 1I)*.

### Functional annotation of genome-wide loci

Of the four genome-wide significant loci *(Figure 1H)* detected for *AERS+*, two directly colocalized with known BMI loci (*FTO* and chr5:104408031-104756336, posterior probability for a single shared causal variant, “PP4” > 0.8, *Methods*). The remaining two loci spanned chr1:72047305-72493356 (index SNP: rs10789340) and chr1:72810145-73611905 (index SNP: rs1852233), with both index SNPs located within *LINC02796*, a long non-coding RNA upstream of *NEGR1*—a gene previously implicated in both BMI and MDD^14,15^. Index variants rs10789340 and rs1852233 were in low LD (r^2^ = −0.22) but remained genome-wide significant after conditional analysis with COJO (*Methods*, *Supplementary Table 5*). To further assess whether these loci were independent of BMI, we conditioned the GWAS on summary statistics for BMI (*Methods*) using mtCOJO^16^. All loci except the locus in *FTO* remained genome-wide significant, with FTO showing the largest attenuation (*Supplementary Table 6*). We corroborated this by directly adjusting for BMI in the *AERS+* analysis in UKBB, observing full attenuation at *FTO* and minimal attenuation at the remaining loci (*Supplementary Table 7*).

We performed eQTL colocalization using single-nucleus eQTL data from human prefrontal cortex tissue to assign candidate effector genes (*Supplementary Table 8*). These results were complemented with SNP-level functional annotations based on evolutionary constraint and candidate cis-regulatory elements (cCREs), functional annotations that are strongly enriched in psychiatric disorder heritability^17,18^. Among *AERS+* loci, chr1:72047305-72493356 colocalized with an eQTL for *LINC02796* in cortical inhibitory neurons (*Figure 2A)*. The index SNP rs10789340, an intronic variant within *LINC02796*, was evolutionarily conserved across primates and was located within a cCRE active in inhibitory neurons (*Supplementary Data*).

**Figure 2.**
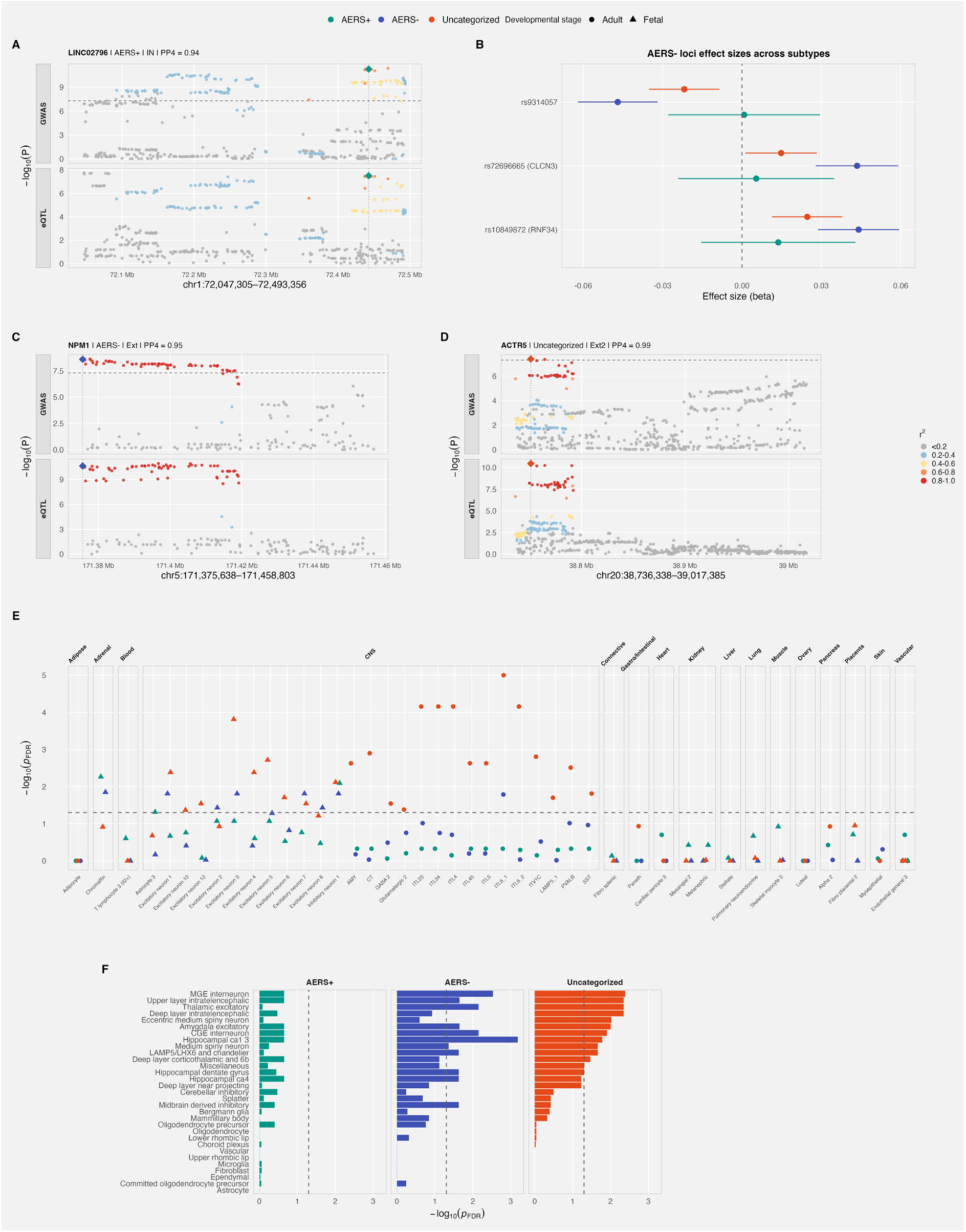
Cell-type, tissue, and eQTL analyses of MDD subtypes. (A) Regional eQTL colocalization plot at an AERS+ locus showing the GWAS signal (top) and the matched eQTL signal (bottom). Points are colored by LD (r²) to the lead variant; the lead GWAS variant is marked as a filled diamond. The dashed line marks P = 5 × 10⁻⁸. Panel title lists the colocalized gene, subtype, eQTL cell type/tissue, and posterior probability of shared causal variant (PP4). (B) SNP effect sizes (β) at AERS− loci (index SNPs: rs9314057, rs72696665, rs10849872) across all three subtypes. Error bars are 95% confidence intervals. (C) Regional eQTL colocalization plot as in (A), for the AERS− locus spanning chromosome 5: 171375638 – 171458803. (D) Regional eQTL colocalization plot as in (A), for the Uncategorized locus spanning chromosome 20: 38736338 – 39017385. (E) Stratified heritability enrichment in open-chromatin regions from fetal and adult cell types. Points are plotted as −log₁₀(P_FDR) for each cell type; shape denotes developmental stage (adult vs. fetal); color denotes subtype. The dashed line marks FDR = 0.05. For readability, cell types are restricted to those that are FDR-significant in at least one subtype or that represent the most significant annotation within a tissue. (F) Stratified heritability enrichment in specifically expressed genes from 31 human brain cell types, shown as −log₁₀(P_FDR) separately for each subtype. The dashed line marks FDR = 0.05.

We detected 10 genome-wide significant loci for *AERS−*, of which two directly colocalized (PP4 > 0.8 in *coloc*, *Method*s, *Supplementary Table 4 & 9*) with metabolic traits). rs9314057, located in *LINC03000*, colocalized with waist circumference loci, with the *AERS−* risk-increasing allele associated with smaller waist circumference. rs10849872, near *RNF34*, colocalized with loci for BMI, fasting glucose, and type 2 diabetes, with the *AERS−* risk allele associated with decreased BMI and reduced type 2 diabetes risk. These loci displayed attenuated effects for *AERS+* (*Figure 2B*). We further detected a genome-wide hit on chromosome 4 for *AERS−* near *CLCN3* which colocalized with both a schizophrenia locus and an eQTL for *CLCN3*, suggesting a shared mechanism through *CLCN3* regulation. eQTL colocalization additionally identified *NPM1* as a candidate effector gene, with an *AERS−*locus colocalizing with increased *NPM1* expression in excitatory neurons (*Figure 2C)*. Functional annotation highlighted rs2071276, a *NPM1* SNP intersecting multiple cCREs in both fetal and adult brain annotations (*Supplementary Data)*. Functional annotation further prioritized rs2806933, a variant downstream of *OLFM4* that intersected multiple cCREs and was constrained in primates (*Supplementary Data*). For the 13 loci associated with the Uncategorized subtype, we assigned four putative effector genes for three distinct loci through eQTL colocalization: *ACTR5* and *PPP1R16B* (in inhibitory and excitatory neurons), lead SNP rs4810212 with *MFAP3L* in inhibitory neurons (*Figure 2D*) and lead SNP rs12650424 with *PKD2* in astrocytes (*Supplementary Figure 5*).

#### Tissue-and cell-type enrichment

We conducted tissue-and cell-type enrichment analysis using stratified LDscore regression (“S-LDSC”) with publicly available scATAC data^19,20^, snRNA-seq data^21^and bulk tissue data^22^ (*Methods, Supplementary Table 10*). We first applied a tissue-agnostic analysis, examining heritability enrichment for AERS subtypes across open-chromatin regions in 256 fetal and adult cell types (*Figure 2E*), broadly representing tissues across the human body. We observed significant enrichment (P_FDR_ < 0.05) exclusively in cell types from the central nervous system - except for fetal adrenal chromaffin cells (*AERS+* and *AERS−)*, a cell type deriving from the neural crest acting like sympathetic neurons during development. This brain-specific pattern was corroborated by a complementary analysis of bulk tissue expression across 37 tissues from GTEx, in which significant heritability enrichment was again confined to central nervous system tissues across all three subtypes (*Supplementary Figure 6*). Given this central nervous system specificity, we further assessed enrichment in genes specifically expressed across 31 major adult human brain cell types^21^. For the *Uncategorized* subtype, we identified cell-types consistent with previous reports of MDD^23^ (*Figure 2F*). For *AERS−* we observed a similar set of cell-types, the strongest association with Hippocampal CA1-3 neurons. For *AERS+*, no significant cell-type associations emerged, although we observed significant enrichment in brain at the tissue level (*Supplementary Figure 6*). We observed no significant enrichment for glial cells in any of the subgroups.

### Subtype-specific genetic links between AERS and metabolic traits

We estimated genetic correlations (*r*_g_) between AERS subtypes and a set of metabolic, cardiovascular, immune, and psychiatric traits using LDSC (*Figure 3A, Supplementary Table 11-12*). Most notably, we observed a clear divergence in *r*_g_ between *AERS+* and *AERS−* with metabolic traits. Across five metabolic-related traits (BMI, waist circumference, metabolic syndrome, type 2 diabetes, random glucose), we observed significantly negative *r*_g_ with *AERS−*, whereas *AERS+* displayed significant positive genetic correlations, for example with BMI (*AERS+*: *r*_g_ = 0.390, se = 0.025, *AERS−*: *r*_g_ = −0.11, se = 0.022) and with metabolic syndrome (*AERS+*: *r*_g_ = 0.362, se = 0.025, *AERS−*: *r*_g_ = −0.068, se = 0.02).

**Figure 3.**
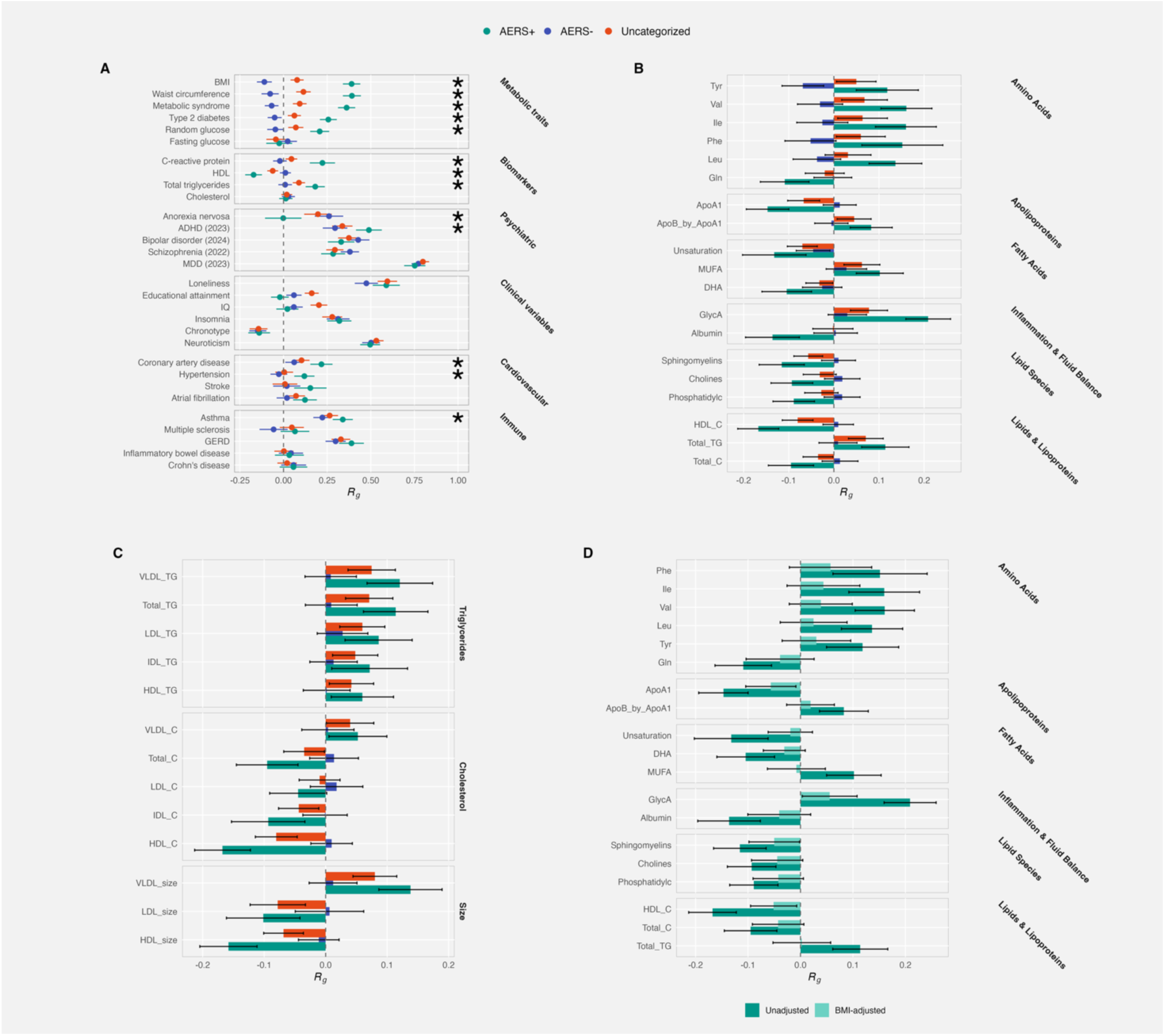
Genetic correlations of MDD subtypes with external traits and biomarkers. (A) Genetic correlations (*r*_g_) of AERS+, AERS−, and Uncategorized MDD with psychiatric, clinical, metabolic, cardiovascular, immune, and biomarker traits. Asterisks mark significantly different genetic correlations with the external trait between AERS+ and AERS−. Significance was determined by block jackknife followed by Bonferroni adjustment for multiple testing. Within each category, traits are ordered by the absolute difference in *r*_g_between AERS+ and AERS−. (B) Genetic correlations with Nightingale NMR biomarkers, restricted to biomarkers at which at least one subtype passes Bonferroni significance (P < 0.05/42). Biomarkers are faceted by metabolite class and ordered within class by |*r*_g_|. Error bars are 95% confidence intervals. (C) Genetic correlations with lipoprotein subclass measures, faceted by measurement type (triglycerides, cholesterol, particle size). Error bars are 95% confidence intervals. (D) AERS+ biomarker genetic correlations before and after BMI adjustment with mtCOJO. Bars show unadjusted (solid) vs. BMI-adjusted (faded) estimates for the same biomarker set as in (B). Error bars are 95% confidence intervals.

Among psychiatric disorders, the genetic correlation between *AERS+* and anorexia nervosa was not significantly different from 0 (*r*_g_ = −0.002, se = 0.053), while *AERS−* correlated moderately (*r*_g_= 0.261, se = 0.042). We additionally observed a significantly stronger genetic correlation between *AERS+* and ADHD (*r*_g_= 0.489, se = 0.038) compared to *AERS−* (*r*_g_ = 0.295, se = 0.03, Δ*r*_g_= −0.194, p =1.95 × 10^−4^). Among cardiovascular disease, we observed significantly stronger genetic correlations between *AERS+* and hypertension (*r*_g_= 0.119, se = 0.03) and coronary heart disease (*r*_g_= 0.217, se = 0.032) compared to *AERS−* (hypertension; *r*_g_ = −0.027, Δ*r*_g_= −0.146, p =1.38 × 10^−4^, coronary heart disease; *r*_g_= 0.061, Δ*r*_g_= −0.156, p =1.78 × 10^−4)^.

Additionally, *AERS+* was negatively correlated with HDL-cholesterol (*r*_g_ = −0.17, se = 0.02) and positively correlated with C-reactive protein (*r*_g_ = 0.22, se = 0.04), neither of which were significant for *AERS−* (C-reactive protein, *r*_g_ = −0.02, se = 0.02; HDL, *r*_g_ = 0.01, se = 0.02). For most traits tested, the Uncategorized subgroup results were consistent with a mix of the two AERS subgroups. The exception was cognitive traits: genetic correlations with educational attainment and IQ were significantly greater in the Uncategorized group than in *AERS+* (Δ*r*_g_= 0.18, p = 3.2 ×10⁻⁸ for EA; Δ*r*_g_= 0.18, p = 8.6 ×10⁻⁶ for IQ) or *AERS−* (Δ*r*_g_= 0.10, p = 7.6 ×10⁻⁴ for EA; Δ*r*_g_= 0.14, p = 6.0 ×10⁻⁵ for IQ).

To characterise this metabolic and inflammatory profile in greater detail, we estimated genetic correlations with 249 circulating metabolite measures from the Nightingale NMR metabolomics platform^24^. *AERS+* showed significant genetic correlations across multiple metabolite classes, whereas *AERS−* showed no significant correlations after adjustment for multiple testing (*Figure 3B-C, Supplementary Table 13*). The pattern of genetic correlations for *AERS+* was consistent with a pro-atherogenic and pro-inflammatory metabolic state. First, *AERS+* showed positive correlations with triglyceride-rich lipoproteins (VLDL triglycerides, *r*_g_ = 0.12, se = 0.027; VLDL particle size, *r*_g_ = 0.14, se = 0.026) and negative correlations with HDL-cholesterol (*r*_g_ = −0.17, se = 0.023) and HDL particle size (*r*_g_ = −0.16, se = 0.024), a profile indicating elevated cardiovascular risk^25^ and impaired reverse cholesterol transport (*Figure 3C*). Second, *AERS+* was associated with higher monounsaturated fat levels, lower fatty acid unsaturation, and elevated branched-chain amino acids (valine, *r*_g_ = 0.16, se = 0.029; isoleucine, *r*_g_ = 0.16, se = 0.034), a profile reflecting established markers of insulin resistance. Glycoprotein acetyls (GlycA), a composite marker of systemic inflammation, showed the strongest association of any metabolite tested (*AERS+*: *r*_g_ = 0.21, se = 0.025, *AERS−*: *r*_g_ = 0.03, se = 0.022, *Figure 3B*). This *AERS+* profile closely resembles the dyslipidaemia and low-grade inflammation characteristic of metabolic syndrome. Conditioning *AERS+* on BMI using mtCOJO (*Methods*) attenuated all metabolite associations (*Figure 3D, Supplementary Table 14*), indicating that these correlations are genetically mediated through adiposity rather than representing an independent metabolic signature. Consistent with this, *AERS+* was positively correlated with measures of overall and central fat distribution (waist-to-hip ratio, visceral and abdominal fat depots; *Supplementary Figure 7)*, but showed substantially attenuated correlation with the corresponding BMI-adjusted traits (e.g. WHR adjusted by BMI, or VAT adjusted by BMI), indicating that the signal is captured by overall adiposity rather than fat distribution.

Furthermore, we derived polygenic scores for each of the AERS subtypes using SBayesRC (*Methods*) and tested their association in a phenome-wide association framework in UKBB, removing overlap with individuals in our GWAS (*Figure 4A, Supplementary Table 15*). We observed stronger associations between PRS-AERS+ with E10 (type 1 diabetes), E11 (type 2 diabetes), E16 (“Other disorders of pancreatic internal secretion”), E66 (“Overweight and obesity”), H36 (“Retinal disorders in diseases classified elsewhere”), and L03 (“Cellulitis and acute lymphangitis”) compared to *AERS−,* despite the substantially smaller sample size in the *AERS+* GWAS.

**Figure 4.**
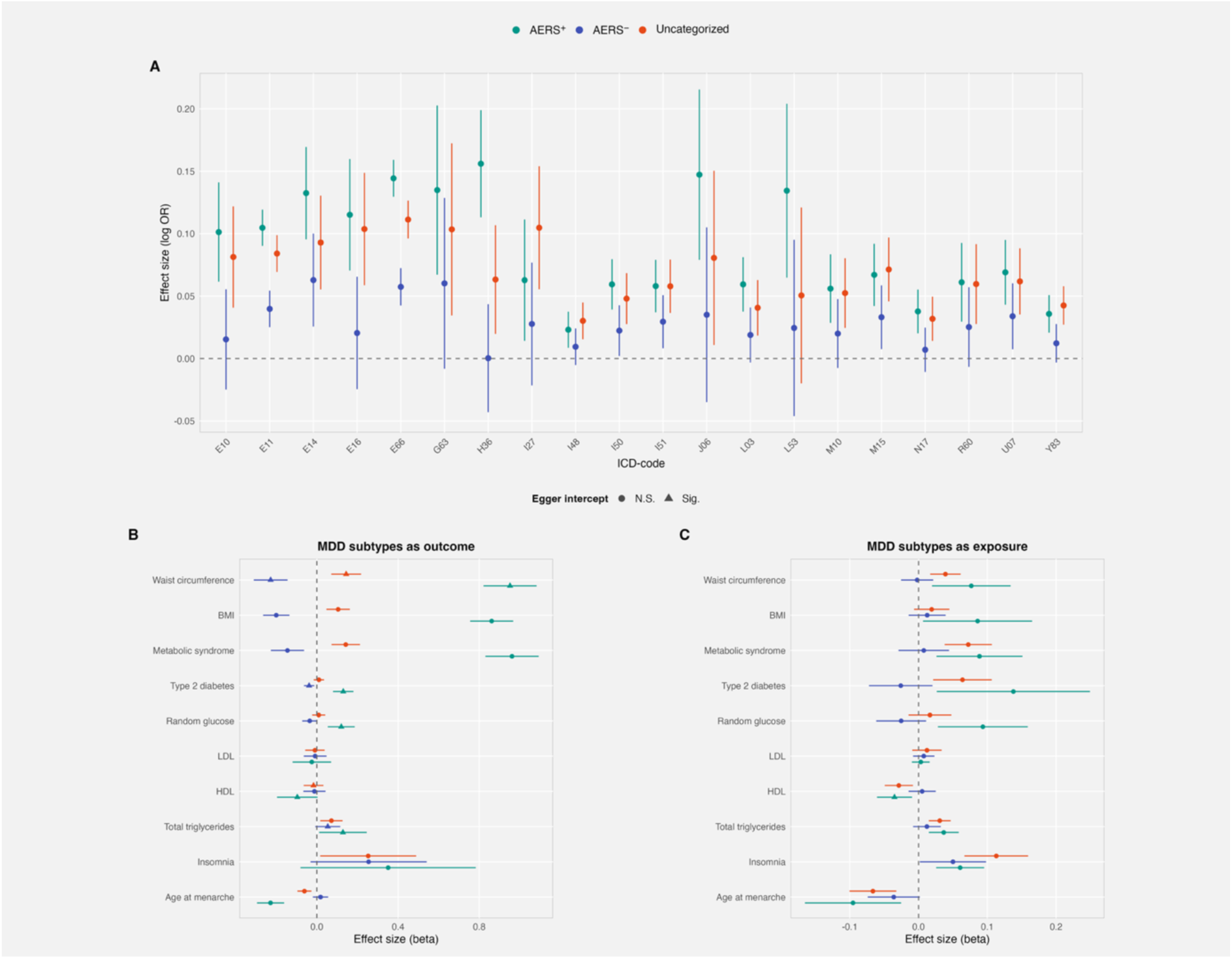
Phenome-wide association and Mendelian randomization analyses. **(A)** PheWAS of AERS+, AERS−, and Uncategorized MDD against ICD-10 outcomes, restricted to the 20 outcomes with the largest AERS+/AERS− effect-size ratio among outcomes where at least one subtype passes Bonferroni significance (P < 0.05/247). Points show effect sizes (log odds ratio); the dashed line marks log OR = 0. ICD-10 codes are ordered alphabetically by chapter on the x-axis. **(B)** Two-sample Mendelian randomization with MDD subtypes as outcomes. Inverse-variance-weighted effect estimates are shown for each exposure–outcome pair. Triangular points indicate exposures with a significant MR-Egger intercept (P < 0.05), signaling potential directional pleiotropy. Error bars are 95% confidence intervals; dashed lines mark β = 0. **(C)** Two-sample Mendelian randomization with AERS subtypes as exposures, displayed as in (B).

#### Putatively causal relationships between AERS subtypes and metabolic traits

We assessed whether obesity- and metabolic-related traits may have a causal influence on the AERS subtypes by performing two-sample Mendelian randomisation, using the inverse-variance weighted estimator as the primary method (*Supplementary Table 16)*. We observed evidence for putatively causal effects of increased genetic liability to metabolic syndrome (*β* = 0.96, se = 0.07, p = 9.23 × 10^−47^), higher waist circumference (*β* = 0.95, se = 0.07, p = 2.46 × 10^−46^), and higher BMI (*β* = 0.86, se = 0.054, p = 2.89 × 10^−58^) on *AERS+*. For *AERS−*, however, we observed significant effects in the opposite direction (metabolic syndrome: *β* = −0.14, se = 0.04, p= 0.0005, waist circumference: *β* = −0.23, se = 0.04, p= 9.07 × 10^−8^, and BMI: *β* = −0.2, se = 0.03, p = 1.57 × 10^−9^, *Figure 4B*). As the MR-Egger intercept for waist circumference was nominally significant (p< 0.05) and borderline significant for BMI (p=0.057), we verified all associations with additional methods robust to pleiotropy, which all indicated effects in the same direction observed with the IVW estimator (*Supplementary Figure 8*). For random glucose, HDL, total triglycerides and type 2 diabetes, we observed evidence of horizontal pleiotropy, and pleiotropy aware estimators did not identify significant results (*Supplementary Figure 8*). Analysis using the AERS subtypes as the exposure indicated bi-directional relationships between *AERS+* and BMI, waist circumference and metabolic syndrome (*Figure 4C*).

### Case-case analysis highlights both metabolic and psychiatric differentiators

We then sought to directly investigate the genetic architecture differentiating *AERS+* and *AERS−* by performing a case-case GWAS, with *AERS+* as cases and *AERS−* as controls, resulting in an effective sample size of 33,039. The case-case GWAS identified four genome-wide significant loci (*FTO*, *ADCY3*, *TMEM18* and *NEGR1, Figure 5A, Supplementary Table 17*), all of which have been extensively described in their association with obesity and adiposity related traits^14^. Adjusting for BMI with mtCOJO led to attenuations in effect sizes for several index SNPs, with the largest attenuation in the *FTO* locus (*Figure 5B, Supplementary Table 18)*. After adjustment, the observed scale heritability was attenuated but remained significant (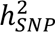 after adjustment = 7.3%, before adjustment: 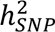 = 13.3%, *Figure 5C*).

**Figure 5.**
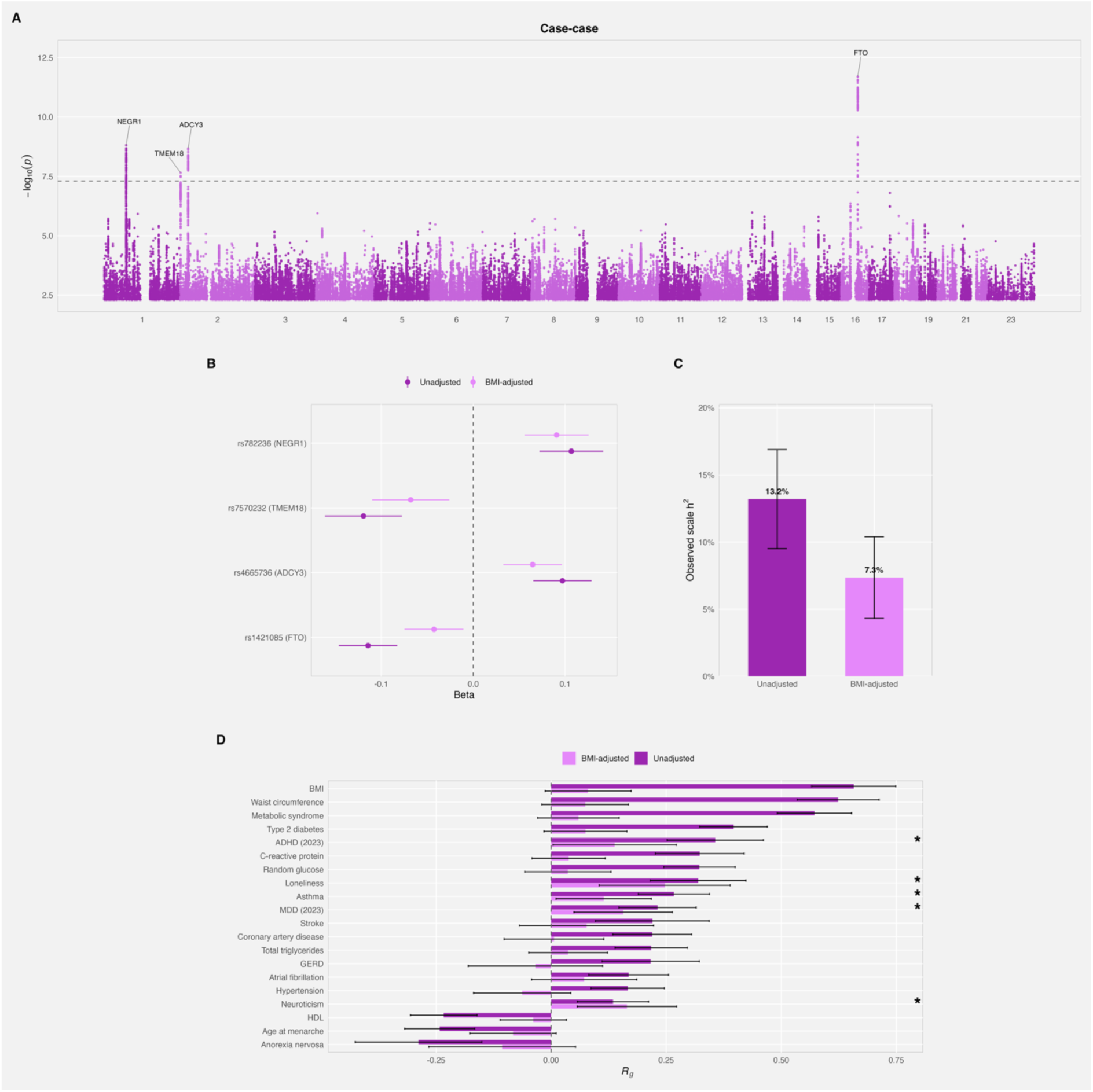
Case-case GWAS contrasting AERS+ vs. AERS−. **(A)** Manhattan plot for the direct case-case GWAS (AERS+ cases vs AERS− cases). Genome-wide significant loci are annotated by nearest gene. The dashed line marks P = 5 × 10⁻⁸. **(B)** SNP effect sizes at case-case lead loci (P < 5 × 10⁻⁸) in the case-case summary statistics, comparing unadjusted vs. BMI-adjusted (mtCOJO) estimates. Error bars are 95% confidence intervals. **(C)** Observed-scale SNP heritability of the case-case phenotype before and after BMI adjustment. Numeric labels give the point estimate as a percentage. Error bars are 95% confidence intervals. **(D)** Genetic correlations between the case-case phenotype and external traits, shown for traits that are Bonferroni-significant in the unadjusted analysis. Solid bars are unadjusted *r*_g_; faded bars are *r*_g_ after BMI adjustment (mtCOJO). Asterisks mark traits that remain nominally significant (P < 0.05) after BMI adjustment. Error bars are 95% confidence intervals.

We computed genetic correlations between the case-case GWAS and a set of traits of interest to better understand the genetic architecture differentiating *AERS+* and *AERS−* (*Figure 5D, Supplementary Table 19*). The case-case GWAS correlated moderately with BMI (*r*_g_= 0.66, se = 0.047). As expected, many genetic correlations with metabolic traits, such as waist circumference, metabolic syndrome and coronary artery disease were substantially attenuated and were no longer significant after adjusting for BMI (*Figure 5D, Supplementary Table 20*). In the BMI-adjusted case-case GWAS, the strongest genetic correlations were with loneliness (*r*_g_= 0.247, se = 0.073, p = 7 ×10^−4^), followed by neuroticism (*r*_g_= 0.165, se = 0.055 p = 2.8 × 10^−3^), and MDD (*r*_g_= 0.156, se = 0.055, p = 0.004). These results suggest that the two subtypes differ not only in metabolic profiles but also in MDD clinical characteristics and patterns of psychosocial traits.

## Discussion

MDD is currently defined as a single diagnostic entity, despite longstanding clinical recognition of both quantitatively and qualitatively different neurovegetative presentations. By leveraging the directionality of appetite/weight and sleep symptoms, we show that these presentations index meaningful differences in genetic architecture between MDD with atypical energy-related symptoms (*AERS+*) and its opposite, MDD with weight loss and hyposomnia (*AERS−*). We analysed the *Uncategorized* group explicitly as it represents the most common subgroup in clinical settings and reflect symptom instability across episodes^3^. The *Uncategorized* group showed a symptom and genetic profile consistent with a mixed AERS related phenotype, with genetic correlations indicating an intermediate position between *AERS+* and *AERS−* groups, except for cognition which showed stronger positive correlations with this group. In addition to the 27 genome-wide significant loci associated with risks for these AERS subtypes, we report several genome-wide significant loci differentiating *AERS+* from *AERS−* subtypes of MDD and a significant 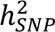 of 13.3% (7.3% when adjusting for BMI).

Two of four genome-wide significant loci for *AERS+* mapped near *NEGR1*, a neuronal growth regulator involved in neurite outgrowth and synaptic connectivity, best known for its association with BMI^14^, but that has been implicated in both feeding behaviour and depression-like phenotypes in mouse models^26^. These two loci were independent of each other and remained significant after adjustment for BMI, demonstrating independence from nearby BMI signals. Notably, this region harbours a complex LD structure with three independent MDD associations and one BMI association in publicly available GWAS^12,14^. Both loci showed directional concordance with *AERS−* and *Uncategorized* samples, but with approximately three-fold larger effect sizes in *AERS+* suggesting that this locus is disproportionately relevant to the *AERS+* subgroup. These results illustrate how symptom-informed subtyping can prioritise genetic signals that are diluted in broad MDD analyses.

Our results highlight metabolic traits as a primary axis of differentiation between *AERS+* and *AERS−* subtypes of MDD. This is supported by directionally distinct genetic correlations with metabolic phenotypes, discordant effect directions in MR, and the case-case analysis directly implicating known BMI genes. Multiple mechanistic interpretations exist. BMI may primarily act as a modifier, shifting the symptom profile of MDD without directly causing the disorder, such that the underlying depression liability is shared across subtypes. Alternatively, metabolic dysfunction may be part of the causal pathway to these specific forms of depression, with chronic systemic inflammation, insulin resistance, or HPA axis dysregulation offering plausible biological mechanisms^7^. These interpretations are not mutually exclusive, and both are consistent with a more fundamental disturbance in energy homeostasis^27^, given that appetite, weight, and sleep are each downstream of central energy-regulatory circuits. Distinguishing between them is challenging with cross-sectional observational data. Without a clear understanding of the specific causal mechanism, simply using BMI as a covariate in the GWAS does not solve this conundrum. If BMI were part of the causal chain leading to *AERS*+, then conditioning on a mediator attenuates the signal it mediates.

However, a purely tautological interpretation that *AERS+* simply captures high-BMI individuals who endorse increased weight and appetite symptoms is not supported by our data. MDD and BMI are bidirectionally associated in longitudinal data^28^, and both our results and previous MR studies support causal effects in both directions^29^. If *AERS+* were reducible to high BMI, we would expect MDD cases matched on BMI to show similar clinical characteristics. Instead, *AERS+* cases show earlier onset and substantially elevated psychiatric comorbidity compared to BMI-matched MDD controls (*Supplementary Note 1; Supplementary Figure 2-3*). Furthermore, the BMI-adjusted case-case analysis retained significant genetic correlations with MDD, neuroticism, and loneliness, indicating that subtype differentiation extends beyond metabolic traits into psychiatric and psychosocial dimensions. In line with this, *AERS+* has been associated with differential antidepressant response and side-effect profiles independent of BMI^30,31^. Taken together, our findings demonstrated clear genetic divergence between these subgroups in both metabolic and depression-related profiles, supporting AERS−related features capturing meaningful subtypes with differential disease biology.

Our findings may also explain why cardiometabolic comorbidity signals have been inconsistent across broad MDD^7,32^: combining subtypes with divergent metabolic genetic liabilities can attenuate or cancel out subtype-specific associations. More generally, syndromic aggregation may inflate apparent polygenicity by pooling partially distinct liability components. The lower polygenicity estimates observed for AERS subtypes (*AERS+*: 1.7%, *AERS−*: 2.9%, *Uncategorized*: 5.3%) are consistent with the hypothesis that high polygenicity in MDD partly reflects etiological mixing rather than uniformly diffuse genetic architecture. From a translational perspective, these results support a precision-psychiatry framing in which metabolic risk factors are more relevant for one depression presentation than another, implying that prevention, stratification, and treatment-response studies should explicitly track symptom directionality rather than relying solely on binary MDD case definitions. The fact that *AERS+* individuals show elevated psychiatric and somatic comorbidity not attributable to their higher BMI additionally suggests the importance of including symptom information in addition to BMI. This distinction may be particularly relevant for metabolic treatments with potential psychiatric effects. For instance, GLP-1 receptor agonists have shown modest effects on overall depression symptoms^33,34^; given the metabolic associations specific to *AERS+*, future research could investigate whether these effects are concentrated within this subtype. More broadly, deep longitudinal phenotyping of clinical and biobank cohorts is essential - specifically, maintaining directionality of weight and sleep items within MDD diagnostic instruments.

Several limitations warrant consideration. First, the genetic analysis was restricted to individuals of European ancestry. We acknowledge that it is particularly important to perform multi-ancestry analyses in AERS subgroups as the relationship between BMI and depression is not consistent across ancestries. For example, an inverse causal relationship has been observed in individuals of East-Asian ancestry when compared to Europeans^35^. However, the availability of large-scale datasets with sufficiently detailed phenotyping, particularly the directionality of neurovegetative symptoms required for subtype definition, remains limited in non-European populations, including in emerging multi-ancestry biobanks such as All of Us. Secondly, the cross-sectional design prevents investigation of symptom stability and causality, and the reliance on retrospective reports introduces potential recall bias. Future studies should prioritize longitudinal designs with granular, symptom-level data to investigate the stability and directionality of weight and sleep dimensions. Third, although we harmonized symptom definitions across cohorts, item wording and ascertainment differed across datasets. This may introduce phenotype misclassification, especially for sleep symptoms where insomnia, early awakening, restless sleep, and reduced sleep duration may not be equivalent constructs. Such misclassification would most likely attenuate subtype differences, but it remains an important limitation. Fourth, treatment exposure was not central to the subtype definition. Antidepressants, antipsychotics, mood stabilizers, and illness chronicity can influence weight, appetite, sleep, and metabolic status, and future cohorts with longitudinal medication data will be needed to separate premorbid subtype biology from treatment-related changes. Finally, biobank and clinical cohort participation may introduce selection and collider effects, particularly when depression severity, BMI, metabolic disease, healthcare contact, and willingness to participate are all related to ascertainment.

In summary, these findings indicate that the directionality of neurovegetative symptoms indexes meaningful genetic heterogeneity within MDD, with metabolic biology emerging as a central, though not exclusive, axis of differentiation. Whether metabolic dysfunction is part of the causal pathway to *AERS+* or primarily modifies its symptomatic presentation remains unresolved, but either interpretation has implications for how depression is studied and eventually treated. More immediately, our results suggest that treating MDD as a genetically homogeneous entity may obscure subtype-specific signals and inflate estimates of polygenicity. Continued efforts to preserve symptom-level detail in both clinical and global biobank settings will be essential for refining these distinctions in future studies.

## Online Methods

### Definition of Lifetime MDD and AERS subtypes

We sought to harmonise definitions of MDD across several cohorts by defining Lifetime MDD based on retrospective worst-episode lifetime depression questionnaires. Lifetime MDD was defined based on the presence of ≥5 of 9 DSM-5 symptoms, moderate to large impact on daily roles, as well as presence of symptoms most of the day, most of the week. Cohort specific details from this procedure are described in the supplementary under “Cohorts”.

We created variables *weight gain* and *weight loss* during the worst depressive episode. If symptoms for both appetite and weight were available, it was sufficient to report either appetite gain or weight gain. However, we required the symptoms to be directionally concordant (for example, individuals reporting weight gain and appetite loss were coded as not reporting weight gain). Reporting “Both gained and lost weight/appetite” did not result in individuals meeting criteria for either *weight_loss* or *weight_gain*. Importantly, weight gain and weight loss were mutually exclusive; an individual could not be coded as reporting both. We created variables *hypersomnia* and *hyposomnia*. The exact phrasing of symptoms varied slightly between cohorts, but “excessive sleep” or “sleeping too much” was coded as *hypersomnia*. Any report of symptoms such as “waking too early”, “restless sleep” or “trouble falling asleep” was coded as *hyposomnia*.

Among individuals meeting criteria for Lifetime MDD we defined AERS+ based on presence of both *weight_gain* and *hypersomnia* during their worst depressive episode. AERS− was defined based on the presence of *weight_loss* and *hyposomnia*. All remaining individuals meeting criteria for Lifetime MDD but not for either AERS− or AERS+ were classified as Uncategorized. Individuals being classified as Uncategorized result from (1) reporting “both gained and lost some weight during the episode” or “stayed about the same or was on a diet” or “Do not know”, resulting in not being classified has having either *weight gain* or *weight loss.* In addition, individuals having only “weight loss” but not hyposomnia or reporting only “weight gain” but not hypersomnia were classified as Uncategorized. Meeting criteria for substance use disorder, bipolar disorder or schizophrenia was considered an exclusion criterion from both controls and cases.

### Genotyping and quality control

The data from each cohort were analyzed separately, restricted to individuals of European ancestry, with principal components and cohort specific covariates used to adjust for confounders and cryptic population stratification. Following GWAS in each cohort we centrally harmonized summary statistics from each cohort and phenotype using tidyGWAS^36^, that applied basic quality control (removing variants with missing values and which were unable to be mapped to dbSNP). We restricted our analysis in each cohort to variants with MAF >= 1%. Insertion and deletions (“indels”) were further removed, but multi-allelic variants were retained if provided from each cohort.

For each summary statistics, we calculated the effective sample size (*N_eff_*) using the formula 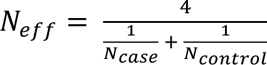, which gives the total sample size comprising equal numbers of cases and controls which has the same power as the sample size of the cohort. In meta-analysis, the SNP-wise *N_eff_*was calculated as the sum of cohort-specific *N_eff_* across all cohorts reporting the SNP, to avoid downward bias in downstream analyses. We conducted fixed-effects inverse-variance weighted meta-analysis across all contributing cohorts within each phenotype, using the meta_analyse function implemented in tidyGWAS. After conducting meta-analysis, we filtered out variants with *N_eff_* < 10,000 and variants with INFO < 0.6.

### Genome-wide loci

We used the clumping algorithm in plink (v.1.9) to identify loci for each trait analysed. The LD reference was the European subset of the 1000 Genomes Project (phase 3, GRCh38, deep WGS)^37^ with parameters: p1 = 5e-8, p2 = 5e-6, r2 = 0.1, and window size of 3000 kb. Overlapping loci and loci within 50 kb of each other were merged using bedtools. We then expanded the loci region by +- 1 MB, and applied GCTA-COJO^38^ to loci with more than 1 index SNP using a LD-reference panel derived from 10,000 randomly selected white British individuals from UK Biobank. Commands --cojo-slct and --cojo-p 5e-8 were used.

### Detecting novel loci

We intersected our genome-wide with loci previously identified in the largest GWAS of MDD to date^12^. For any locus not intersecting this variant set, we further queried the NHGRI-EBI GWAS catalog (https://www.ebi.ac.uk/gwas/home). Loci not detected in the most recent MDD GWAS, and without any related MDD phenotype reported in GWAS catalog was classified as novel.

### Case-Case analysis

We directly conducted case-case GWAS within each contributing cohort and used *AERS−*individuals as control and *AERS+* as cases, using identical covariates and GWAS software as for the main GWAS of *AERS* subtypes compared to controls. Summary statistics were processed in an identical manner as described above. We excluded the *PGC2* cohort in the case-case analysis as the sample size was deemed too small in the individual *PGC2* cohorts for case-case analysis.

### BMI summary statistics

We meta-analysed summary statistics from the UK Biobank and FinnGEN, acquired from the Neale Lab round 2 GWAS and the FinnGen R12 freeze:

- https://storage.googleapis.com/finngen-public-data-r12/summary_stats/release/finngen_R12_BMI_IRN.gz
- https://broad-ukb-sumstats-us-east-1.s3.amazonaws.com/round2/additive-tsvs/21001_irnt.gwas.imputed_v3.both_sexes.tsv.bgz

Summary statistics were downloaded and processed with tidyGWAS, followed by fixed-effects meta-analysis implemented in tidyGWAS::meta_analyse.

### Conditional analysis using mtCOJO

To remove the genetic contribution of body mass index (BMI) from our *AERS+* summary statistics, we performed a conditional analysis using multi-trait-based conditional and joint analysis (mtCOJO), as implemented in GCTA^16^. Analyses were conducted using default parameters, with LD scores derived from 1000 Genomes European subset. The resulting conditional *AERS+* summary statistics and case-case analysis, adjusted for BMI, were used in downstream analyses.

### Phenotypic adjustment for BMI in the UK Biobank

We explored the impact of directly adjusting for phenotypic BMI in the analysis of *AERS+* in UK Biobank. We used the same definition of *AERS+* as for the main GWAS analysis. We conducted GWAS using plink2 with the --glm command and compared the impact of including BMI (BMI at recruitment, field 21001) as covariate, in addition to sex, age and the first 10 principal components.

### Heritability and genetic correlations

We used the LDscore regression^39^ implemented in the R package ldsR^36^ to estimate SNP heritability and genetic correlations. We used publicly available LDscores from the European subset of 1000G. Sample size was set as *N_eff_*. To convert estimates to the liability scale we estimated the proportion of each subtype (out of all MDD individuals) within each cohort and averaged the estimate and multiplied by 0.15 (population prevalence for all MDD). This results in population prevalences of 1.9% for AERS+, 4.7% for AERS− and 8.3% for Uncategorized. To avoid overstating liability-scale heritabilities, we adopted conservative rounded estimates of 1.5% for AERS+, 5% for AERS− and 8.5% for Uncategorized.

We selected summary statistics from a range of psychiatric, cardiometabolic, lifestyle and health determinants to conduct genetic correlations (*Supplementary Table 11*). In addition, we downloaded summary statistics for 249 Nightingale biomarkers from a meta-analysis^24^ of the full UK biobank with Estonia Biobank and used summary statistics restricting to European ancestry (N = 599,249). Summary statistics were processed with tidyGWAS and restricted to MAF > 1%. Summary statistics were subsequently used to estimate genetic correlations using LD score regression, in the R package ldsR. To test whether estimates of *r*_g_ differed between AERS subtypes with external traits, we used a block jackknife procedure over 200 blocks of contiguous SNPs, as previously described^40^. Briefly, per-block delete values for genetic covariance and heritability were used to construct per-block estimates of *r*_g_ for each pair of traits. Jackknife pseudo values were then computed for the difference between the two genetic correlations and used to derive a standard error and two-sided p-value under the null hypothesis of no difference. This approach accounts for the non-independence of estimates arising from shared SNPs and LD structure, which is not captured by the asymptotic standard errors reported by LD score regression. We used FDR for multiple-test adjustment. We calculated and visualized the genetic distance between *AERS+*, *AERS−*, Uncategorized and controls by extending the transformations from GDIS to allow for three subtypes^13^. Specifically, genetic correlations were converted to angles using the inverse cosine, and heritability was converted to genetic distance by taking its square root when on the 50/50 observed liability scale.

### Estimation of polygenicity

We used SBayesS^41^ to derive estimates for polygenicity, defined as the proportion of SNPs exhibiting nonzero effects. We used the publicly available LD matrix (‘ukbEURu_imp_v3_HM3_n50k.chisq10’) derived from UK Biobank data. To assess convergence and ensure robustness of parameter estimates, four parallel Markov chains were executed (–num-chains 4), each comprising 25,000 total iterations (–chain-length 25000), with an initial 5,000 iterations designated as burn-in (–burn-in 5000). We considered a Gelman-Rubin scale-reduction factor of < 1.2 as evidence of algorithmic convergence.

### Cell-type enrichment analyses of SNP-based heritability

#### Open-chromatin region enrichment analysis

To identify cell types showing enrichment of SNP-based heritability, we performed stratified LD Score regression (LDSC), adjusting for the standard 53 baseline annotations. We first downloaded BED files defining open-chromatin regions for 256 human cell types derived from fetal and adult tissuess^19,20^. Genomic coordinates were lifted over to GRCh37 using LiftOver. Cell-type annotations were defined by combining all open-chromatin regions per cell type into a single annotation, and corresponding LD scores were calculated. For the analysis of enrichment in adult brain cell-types we further adjusted for a single annotation containing all open chromatin regions across all adult human brain cell-types. Heritability enrichment was estimated individually per cell type using the R package ldsR, and results were corrected for multiple testing using a false discovery rate (FDR < 0.05) per phenotype.

#### Single-nucleus RNA-seq enrichment analysis

We assessed tissue- and cell-type-specific heritability enrichment using two complementary datasets, in both cases defining annotations by selecting genes ranked within the top 10% by their proportion of total expression within each tissue or cell type^42^. To capture potential associations between AERS subtypes and gene expression across the body, we first examined bulk RNA-sequencing data from GTEx v8^22^ spanning 37 tissues, with annotation BED files obtained from Bryois et al. 2020^43^. We then performed a higher-resolution analysis of brain cell types using single-nucleus RNA-sequencing (snRNA-seq) data from the Human Brain Atlas, comprising 3.37 million nuclei sampled from 106 brain regions^21^; following standard quality control, nuclei were organized into a hierarchical taxonomy of 31 superclusters and 461 fine-grained cell-type clusters (approximately 1,300 genes per cluster), with pre-computed LD scores obtained from Yao et al^23^. Stratified LDSC^44^ was implemented using the R package ldsR, and statistical significance was determined using FDR correction (FDR < 0.05) per phenotype.

### Functional annotation of single variants

We acquired SNP-level functional annotations from previously published sources; variant constraint^17^ and snATAC-seq open-chromatin regions^19,20^. Both annotations have been demonstrated to be massively enriched in psychiatric SNP heritability^18^. We use the Bioconductor package GenomicRanges to convert open-chromatin peaks to SNP-level annotations. To facilitate interpretation, we collapsed the epigenetic data into three meta-annotations: (1) constrained variants, based on mammalian or primate constraint in the Zoonomia dataset; (2) a union of snATAC-seq open-chromatin regions across excitatory and inhibitory fetal neurons; and (3) a union of snATAC-seq open-chromatin regions across excitatory and inhibitory adult neurons. We intersected GWAS summary statistics with these SNP-level functional annotations and manually examined each locus.

### Colocalization with eQTL data and other traits

We performed eQTL colocalization analysis using single-nucleus eQTL summary statistics from the singleBrain meta-analysis^45^. For each genome-wide significant GWAS locus (defined as the lead SNP ± the clumping window; the MHC region, chr6:27–34 Mb, was excluded), we tested colocalization against every cell-type × gene pair available in the fixed-effects eQTL meta-analysis using coloc.abf from the coloc R package^46^, with default priors (p1 = p2 = 1 × 10⁻⁴, p12 = 1 × 10⁻⁵). GWAS variants were restricted to biallelic SNPs with an assigned rsID; missing effect allele frequencies were imputed from 1000 Genomes European allele frequencies^37^, and the effective sample size was used as N. From the eQTL dataset, variants with β = 0 were removed, and a given gene × cell-type pair was tested only if at least one eQTL SNP reached P < 5 × 10^−6^. Colocalization was evaluated on the intersection of SNPs present in both datasets, and a signal was declared colocalized when PP.H4 ≥ 0.80. To ensure adequate overlap, we further required that at least 50% of SNPs in the AERS subtype summary statistics at a given locus were also present in the eQTL summary statistics, and at least 100 SNPs in the intersection. Loci failing this threshold were excluded. Colocalization with other GWAS traits was performed similarly, but we required at least one variant with P < 5 × 10^−8^ to perform colocalization.

### Mendelian randomisation

We performed two sample Mendelian Randomisation (MR) using the “TwoSampleMR” R package^47^. The assumptions of MR have been described in detail elsewhere^48^. We processed summary statistics for traits of interest using tidyGWAS. We defined instruments for each trait using the plink “clumping” approach described above but applied a stricter threshold of r^2^ < 0.01 and p < 5e-08. For the analysis using AERS subtypes as exposures, we relaxed the threshold for instruments to and p < 5e-06, given the smaller number of variants detected at p < 5e-08. Data was harmonized with the outcome and exposure summary statistics using the *TwoSampleMR::harmonise_data* function. We used the inverse-variance weighted (IVW) estimator as our primary method but applied several checks to guard against violations of MR assumptions. We used the MR-Egger method to examine indications of pleiotropy. An association was considered putatively causal if; (1) MR-Egger intercept was non-significant, the IVW estimate was significant after multiple testing correction with FDR and same direction of effect for both MR-Egger and weighted median estimators or (2), the MR-Egger intercept was significant *but* the weighted median and weighed mode estimator was significant, with same direction of effect as the MR-Egger estimate.

### PheWAS

We derived polygenic scores using the SBayesRC method^49^ using standard parameters. We applied ‘SBayesRC::tidy()’ followed by ‘SBayesRC::impute()’ prior to running ‘SBayesRC::sbayesrc()’. We used a publicly available LDmatrix and set of 96 functional annotations available at https://gctbhub.cloud.edu.au/data/SBayesRC/resources/v2.0/. Polygenic scores were calculated in the UK biobank dataset using the PLINK 2 --score command followed by standardisation to mean = 0, sd = 1. We derived a set of individuals in UK Biobank which were not overlapping with any of the cohorts included in the full meta-analysis. To perform this analysis, we were required to remove the GLAD+UKBB analysis from our AERS subtype meta-analysis and used only our set of UKBB samples for GWAS. To ensure no sample overlap between GWAS and PheWAS samples, we removed all individuals who were in our GWAS and further restricted to individuals without relatives in the UK biobank, using the *in.kinship.table* (field 531) variable, and to individuals of European descent. We defined binary outcome variables based on the presence of a unique ICD-codes in data field 41270 in UKBB. ICD codes were binned based on first two digits (“F432” = “F42”). For each polygenic score (Uncategorized, *AERS+*, and *AERS−*), we fit a separate logistic regression adjusting for sex and the first seven principal components.

## Code availability

Analytical code used is available at Github: https://github.com/Ararder/AERS-GWAS

## Data availability

Summary statistics for *AERS+*, *AERS−*, *Uncategorized* and the case-case analysis are publicly available at https://zenodo.org/records/20610434

## Author contributions

S.M. and Y.L. proposed the study.

A.H. and S.M. coordinated the project.

The core analytical team consisted of A.H., S.M.

The core writing group consisted of A.H., S.M., P.T., Y.L.

J.B, A.B.T, W.J.P, J.A.P, C.F contributed to the data analysis and interpretation of results.

A.H, S.M, T.G, R.K, G.P.M, A.A.G, S.B, E.A, F.H, R.W, J.G.T, S.K contributed to the data analysis of an individual study.

Y.L, P.A.T, I.B.H, N.G.M, B.W.J.H.P, S.E.M, B.L.M, A.S.F.K, D.I.B, Y.M,K.L, T.C.E, C.M.L, G.B, D.F.L, J.B.P, J.S, N.R.W contributed to the data or obtained funding. P.T., Y.L. jointly supervised the project.

All authors commented on the manuscript and approved the final version.

## Acknowledgements

We thank Patrick F. Sullivan and Cynthia Bulik for their input on the manuscript. Y.L was supported by the European Research Council (Grant Agreement ID: 101042183), the US National Institute of Mental Health (NIMH) R01 MH123724, and the Swedish Research Council (Vetenskapsrådet; Award 2021-02615). S.E.M is supported by NHMRC grant 2025674. B.M is supported by National Health and Medical Research Council (NHMRC) of Australia grant 1086683. A.S.F.K is supported by a Wellcome Early Career Award (Grant Ref: 227063/Z/23/Z). S.M was supported by Medical Research Council (MR/N013166/1) funding. To create the BIONIC consortium, we are grateful for funding from the Biobanking and Biomolecular Resources Research Infrastructure (BBMRI-NL: 184.021.007; 184.033.111) (D.I.B. and B.W.J.H.P.). This work was supported by the KNAW Academy Professor Award (PAH/6635) (to D.I.B. and used to support F.H.), a NARSAD Young Investigator Grant, and the National Institute of Mental Health (NIMH) grant R01MH125902 (H.M.vL.). Y.M is supported by Amsterdam UMC (Starter Grant Ronde 2), Amsterdam Neuroscience (PoC Funding 2024–2026), and the ImmunoMIND consortium, funded by UK Research and Innovation as part of the UK National Mental Health Platform. Research in EstBB was supported by the Estonian Research Council grants PSG615 and Estonian Center of Genomics/Roadmap II grant TT17, and the Estonian Centre of Excellence for Well-Being Sciences, funded by grant TK218 from the Estonian Ministry of Education and Research. The TEDS data collection included in these analyses was funded by a programme grant to Prof. Thalia Eley from the UK Medical Research Council (MR/V012878/1). This work was also supported by the National Institute for Health and Care Research (NIHR) BioResource [RG94028, RG85445], NIHR Biomedical Research Centre [IS-BRC-1215-20018], HSC R&D Division, Public Health Agency [COM/5516/18], MRC Mental Health Data Pathfinder Award (MC_PC_17217), and the National Centre for Mental Health through Health and Care Research Wales. N.M was supported by the National Health and Medical Research Council (NHMRC) of Australia. J.P was supported by the National Institutes for Health (NIH). S.H was supported by the National Institute of Mental Heatlth (NIMH). J.B. was supported by Hjärt-Lungfonden (the Swedish Heart-Lung Foundation; grant number [20251020]). We acknowledge The Swedish Twin Registry for access to data. The Swedish Twin Registry is managed by Karolinska Institutet and receives funding through the Swedish Research Council under grant no. 2021-00180. The computations were enabled by resources provided by the National Academic Infrastructure for Supercomputing in Sweden (NAISS), partially funded by the Swedish Research Council (grant number: 2021–02615). Data analysis in the Estonian Biobank was carried out in part in the High-Performance Computing Center of University of Tartu. Estonian Biobank research team: Andres Metspalu, Lili Milani, Tõnu Esko, Reedik Mägi, Mari Nelis and Georgi Hudjashov. AGDS was primarily funded by the National Health and Medical Research Council (NHMRC) of Australia Grant No. 1086683, and the QSkin study was funded by the NHMRC (Grant Numbers 1185416, 1063061 and 1073898). We thank everyone who contributed to the conception, implementation, media campaign and data cleaning of the AGDS project, including Richard Parker, Simone Cross, and Lenore Sullivan. Thank you to Scott Gordon for carrying out the imputation and quality control of the AGDS and QSkin data and Penelope Lind for contributing to the management of the AGDS database. This work was supported by NHMRC Investigator Grants to Jackson G Thorp (2027002); Brittany L Mitchel (2017176); Sarah E Medland (1172917); Nicholas G Martin (1172990); Naomi R Wray (1173790); David C. Whiteman (2026567) and Ian B Hickie (2016346). Generation Scotland received core funding from the Chief Scientist Office of the Scottish Government Health Directorate (Grant No. CZD/16/6) and the Scottish Funding Council (Grant No. HR03006). Genotyping and DNA methylation analysis of the Generation Scotland: Scottish Family Health Study samples were carried out by the Genetics Core Laboratory at the Wellcome Trust Clinical Research Facility, Edinburgh, Scotland, and were funded by the U.K. MRC and the Wellcome Trust (Wellcome Trust Strategic Award “Stratifying Resilience and Depression Longitudinally”) (Reference No. 104036/Z/14/Z). This work has made use of the resources provided by the Edinburgh Compute and Data Facility (ECDF) and MRC equipment award (Award No. MC_PC_MR/X013677/1). We thank all the families who took part, the general practitioners and the Scottish School of Primary Care for their help in recruiting them, and the whole Generation Scotland team, which includes interviewers, computer and laboratory technicians, clerical workers, research scientists, volunteers, managers, receptionists, health care assistants, and nurses. The BIObanks Netherlands Internet Collective (BIONIC) was made possible through the work and trust of the collaborating cohorts and their participants. We are grateful to all individuals who participated in this research, and to the many colleagues who contributed to this project and its constituent studies. We gratefully acknowledge the ongoing contribution of the participants in the Twins Early Development Study (TEDS) and their families. We thank NIHR BioResource volunteers for their participation and gratefully acknowledge NIHR BioResource centres, NHS Trusts, and staff for their contribution. We thank the National Institute for Health and Care Research, NHS Blood and Transplant, and Health Data Research UK as part of the Digital Innovation Hub Programme. The views expressed are those of the author(s) and not necessarily those of the NHS, the NIHR, or the Department of Health and Social Care. We gratefully acknowledge the participation of the NIHR BioResource Centre Maudsley, Biomedical Research Centre at South London and Maudsley NHS Foundation Trust, and King’s College London volunteers, and thank the staff for their help with volunteer recruitment. We thank the NIHR Biomedical Research Centre at South London and the Maudsley NHS Foundation Trust and King’s College London for funding. This study represents independent research supported by the NIHR Biomedical Research Centre BioResource at South London and Maudsley NHS Foundation Trust and King’s College London.

## Conflicts of interest

IBH is Professor of Psychiatry and the Co-Director of Health and Policy, Brain and Mind Centre, University of Sydney. He has led major public health and health service developments in Australia, particularly focusing on early intervention for young people with depression, suicidal thoughts and behaviours and complex mood disorders. He is active in the development through codesign, implementation and continuous evaluation of new health information and personal monitoring technologies to drive highly-personalised and measurement-based care. He holds a 3.2% equity share in Innowell Pty Ltd that is focused on digital transformation of mental health services. The remaining authors declare no conflicts of interests.

## Notes

### Author Declarations

This study was approved by the Swedish ethical review authority (Dnr 2023-03073), and each cohort was approved by the relevant institutional review boards.

