## Supplementary text for "Atypical energy-related symptoms define biologically distinct subtypes of major depressive disorder"

### Supplementary Note

AERS+ individuals had substantially higher average BMI than AERS-, Uncategorized, and control individuals (Supplementary Figure 1), consistent with previous studies of weight/appetite-based MDD subtypes^1,2^. Given this difference, and the mechanical link between AERS+ criteria and BMI (weight/appetite gain during the index episode), we asked whether AERS subtyping captures meaningful distinctions or whether it indirectly stratifies MDD cases on BMI. Dahl & Zaitlen (2020)^3^ illustrate this concern with a hypothetical GWAS of diabetes stratified by hair colour: subtype-specific signals would emerge, but they would reflect pigmentation genetics rather than diabetes aetiology.

Unlike hair colour and diabetes, however, BMI and MDD are not etiologically independent. Epidemiological studies support a bidirectional association, elevated BMI predicts subsequent MDD and depressive episodes predict subsequent weight gain, with a U-shaped

risk curve where both underweight and overweight is associated with MDD and stronger effects in females^4,5^. Mendelian randomisation analyses are consistent with bidirectional causation^6^. BMI is therefore not a neutral stratifier in the Dahl & Zaitlen sense, but rather a trait entangled with MDD pathophysiology. Nevertheless, the underlying concern that an AERS+ subtype signal may be dominated by BMI-associated biology rather than depression-specific biology remains worth addressing empirically.

To do so, we used UK Biobank data from all MHQ1 or MHQ2 participants (N = 217,124). Lifetime MDD and AERS subtypes were defined as in our GWAS, without ancestry or genotype-based exclusions (lifetime MDD = 53,917; AERS- = 22,395; AERS+ = 3,280; Uncategorised = 28,242; controls = 163,062). We then matched each AERS+ individual 1:1 on BMI and sex to all non AERS+ MDD cases using the R package MatchIt and performed the analogous matching for AERS- (all non AERS- MDD cases).

Relative to both the BMI-matched MDD comparison group and the other AERS subtypes, AERS+ individuals showed earlier age at onset, higher rates of recurrence, and a greater proportion reporting substantial functional impairment indicating that the clinical profile of AERS+ is not captured by BMI and sex alone.

| Group | n | Female (%) | BMI | Age at onset (y) | Recurrent (%) | Substantial functional impairment (%) |
| --- | --- | --- | --- | --- | --- | --- |
| AERS+ | 3,280 | 74.9 | 30.6 | 31.6 | 76.7 | 67.9 |
| BMI-matched to AERS+ | 3,280 | 74.9 | 30.6 | 35.8 | 60.3 | 52.6 |
| AERS- | 22,395 | 73.5 | 26.3 | 35.8 | 59.1 | 54.6 |
| BMI-matched to AERS- | 21,386 | 73.0 | 26.5 | 34.5 | 60.9 | 50.7 |
| Uncategorised | 28,242 | 64.0 | 27.5 | 35.2 | 59.7 | 49.8 |
| Lifetime MDD | 53,917 | 68.6 | 27.2 | 35.3 | 60.5 | 52.9 |

**Table 1**:MDD self-reported characteristics for AERS+, AERS- and Uncategorised in UK biobank. Comparison groups contain propensity matched MDD cases on BMI and sex. The table shows % female, mean BMI, mean age at onset (self-reported), % reporting 2 or more episodes, and % reporting substantial functional impairment.

We next examined comorbidity using logistic regression for a panel of psychiatric and somatic conditions (ascertained from self-report and ICD-coded registry data), adjusting for age, sex, Townsend index, assessment centre, and smoking status. AERS+ was associated with substantially elevated risk of anxiety, bipolar disorder, binge eating, and OCD, among others (*Supplementary Figure 2-3*). Because BMI was matched between the AERS+ and comparison groups by design, these associations cannot be attributed to BMI differences. Together, these analyses show that AERS+ differs from other MDD cases in both clinical course and comorbidity profile, and that these differences are not explained by elevated BMI.

### Supplementary Figures

##### Supplementary Figure 1

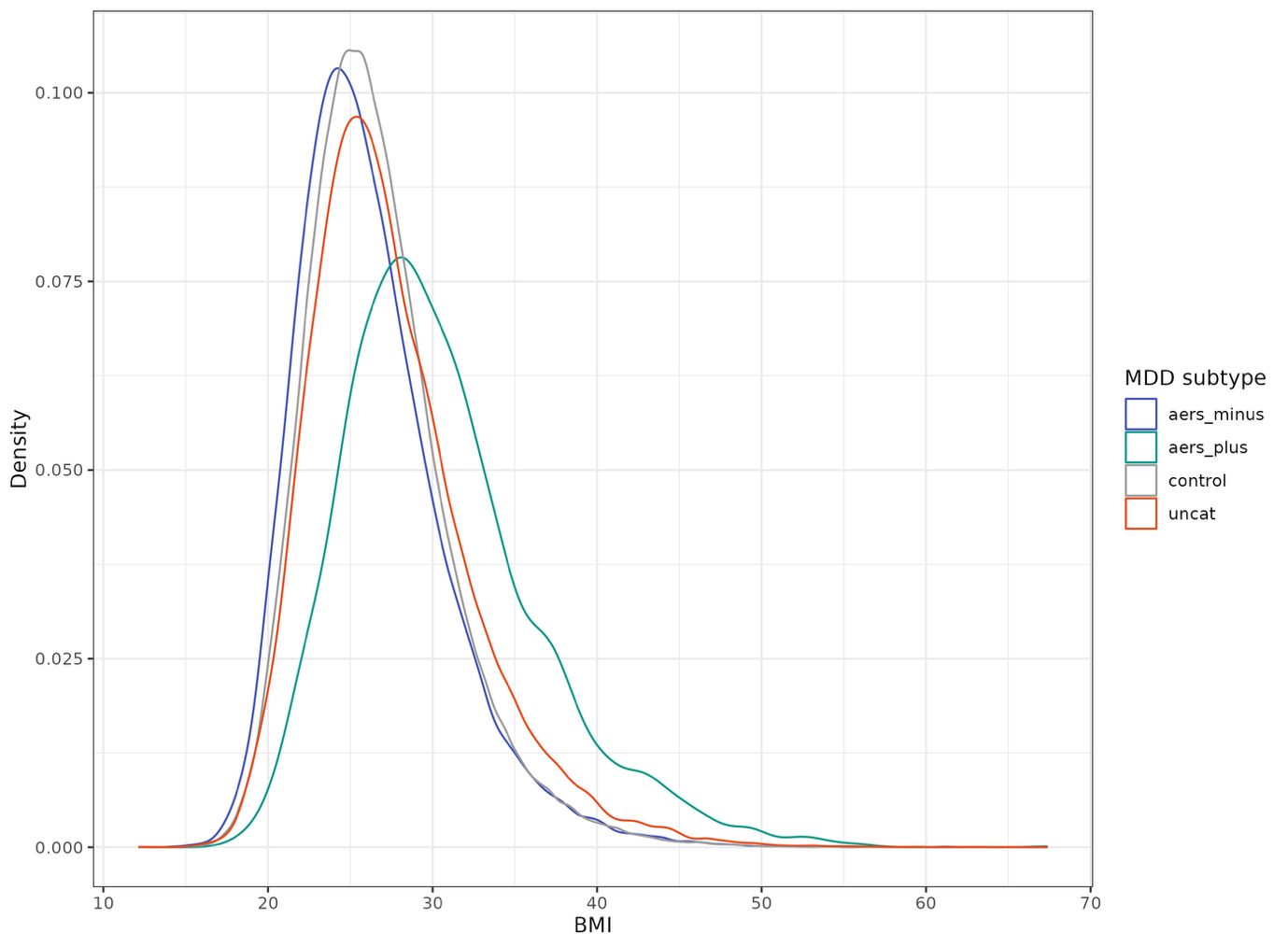

**Supplementary Figure 1**. Distribution of BMI in the UK Biobank MHQ1 and MHQ2 sample (N = 217,124). Densities are shown separately for lifetime MDD cases stratified by AERS subtype (AERS+, AERS-, Uncategorized) and controls.

##### Supplementary Figure 2

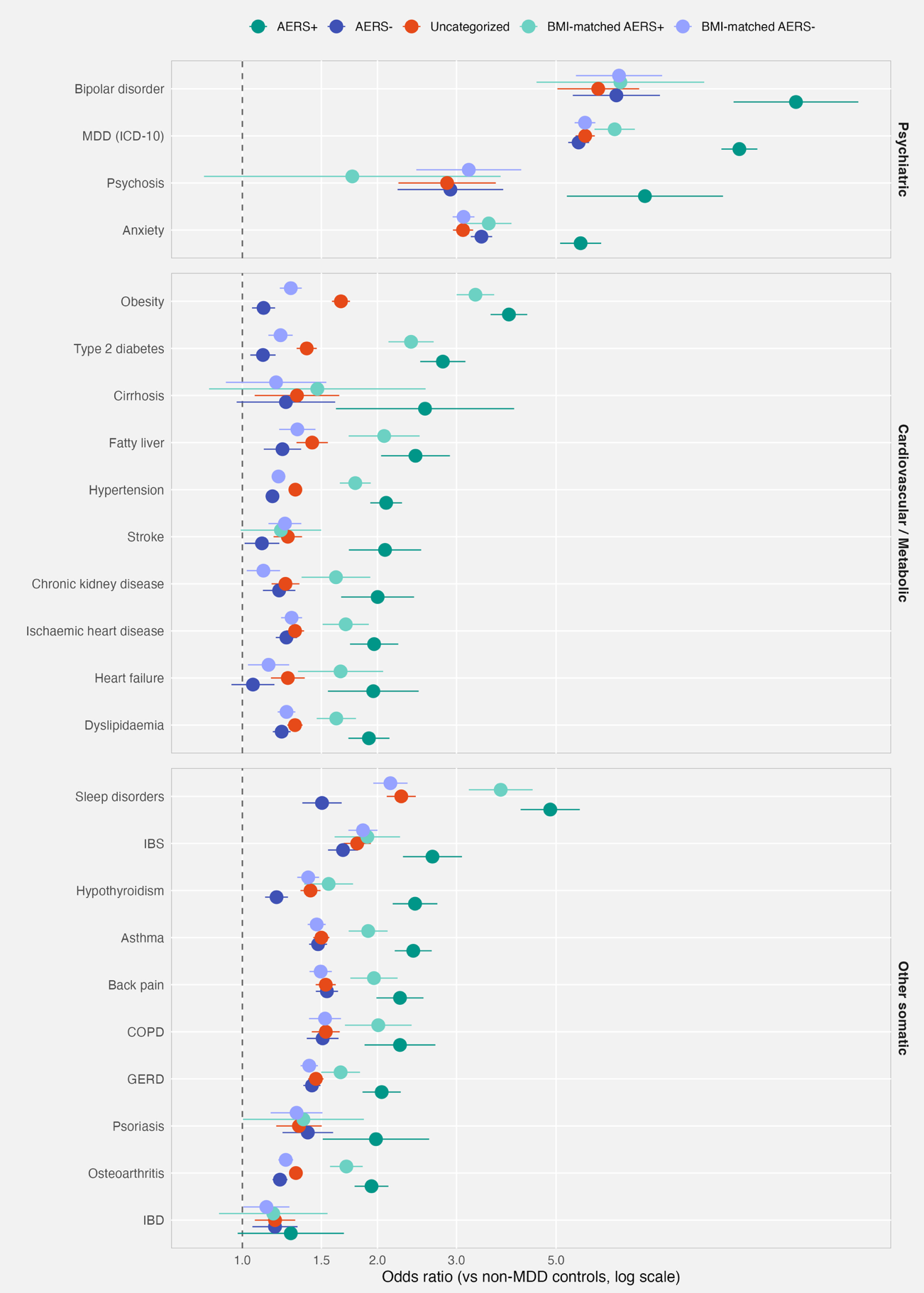

**Figure 2:** ICD-coded comorbidity odds ratios (ORs) comparing AERS+, AERS-, and Uncategorized MDD to BMI-matched MDD comparison groups in UK Biobank. ORs were estimated by logistic regression adjusting for age, sex, Townsend index, assessment centre, and smoking status. Error bars are 95% confidence intervals; the dashed line marks OR = 1.

##### Supplementary Figure 3

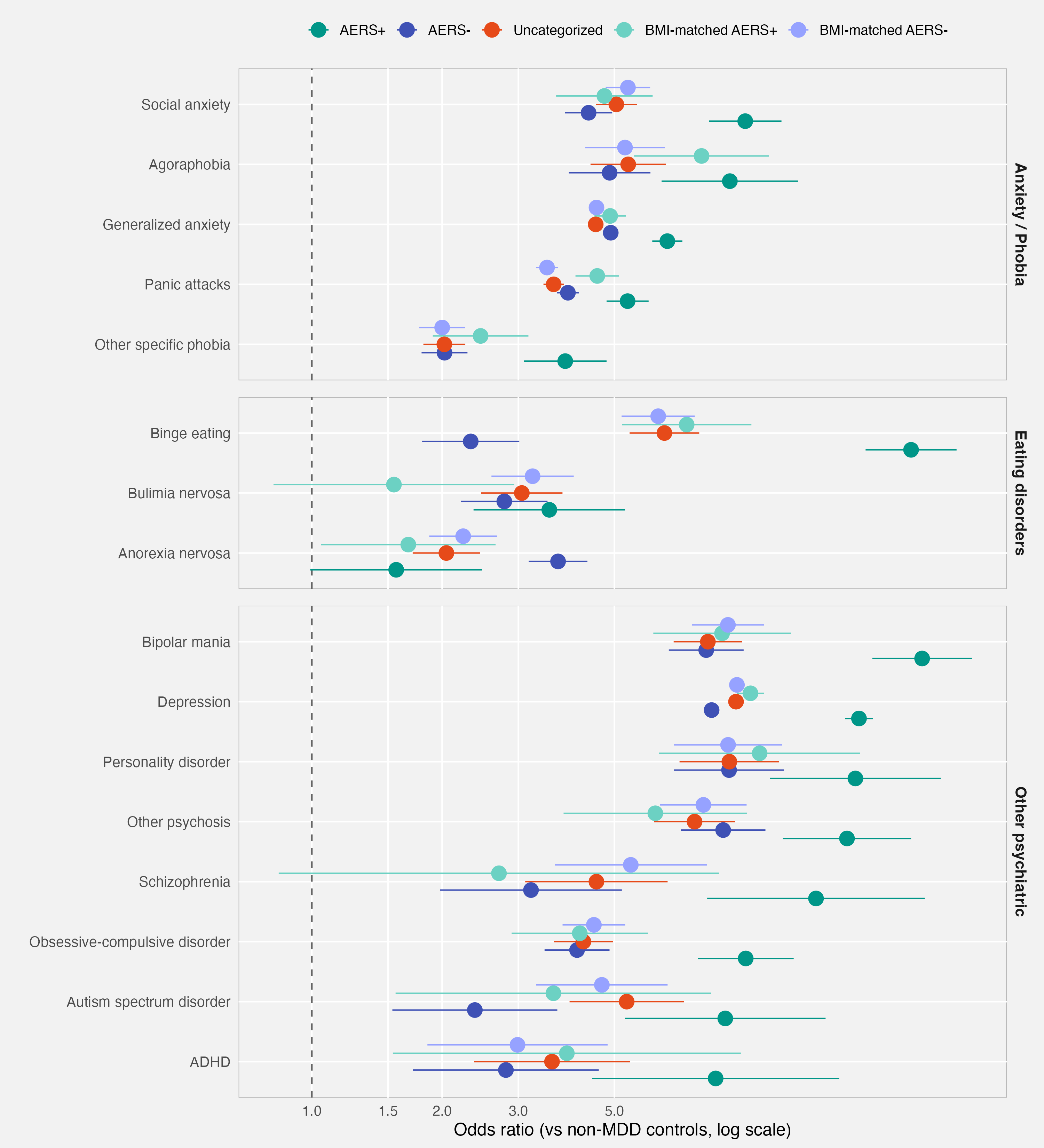

**Figure 3**: Self-report comorbidity ORs for psychiatric conditions, estimated as in Supplementary Figure 2 but using UK Biobank self-report data. Error bars are 95% confidence intervals; the dashed line marks OR = 1.

##### Supplementary Figure 4

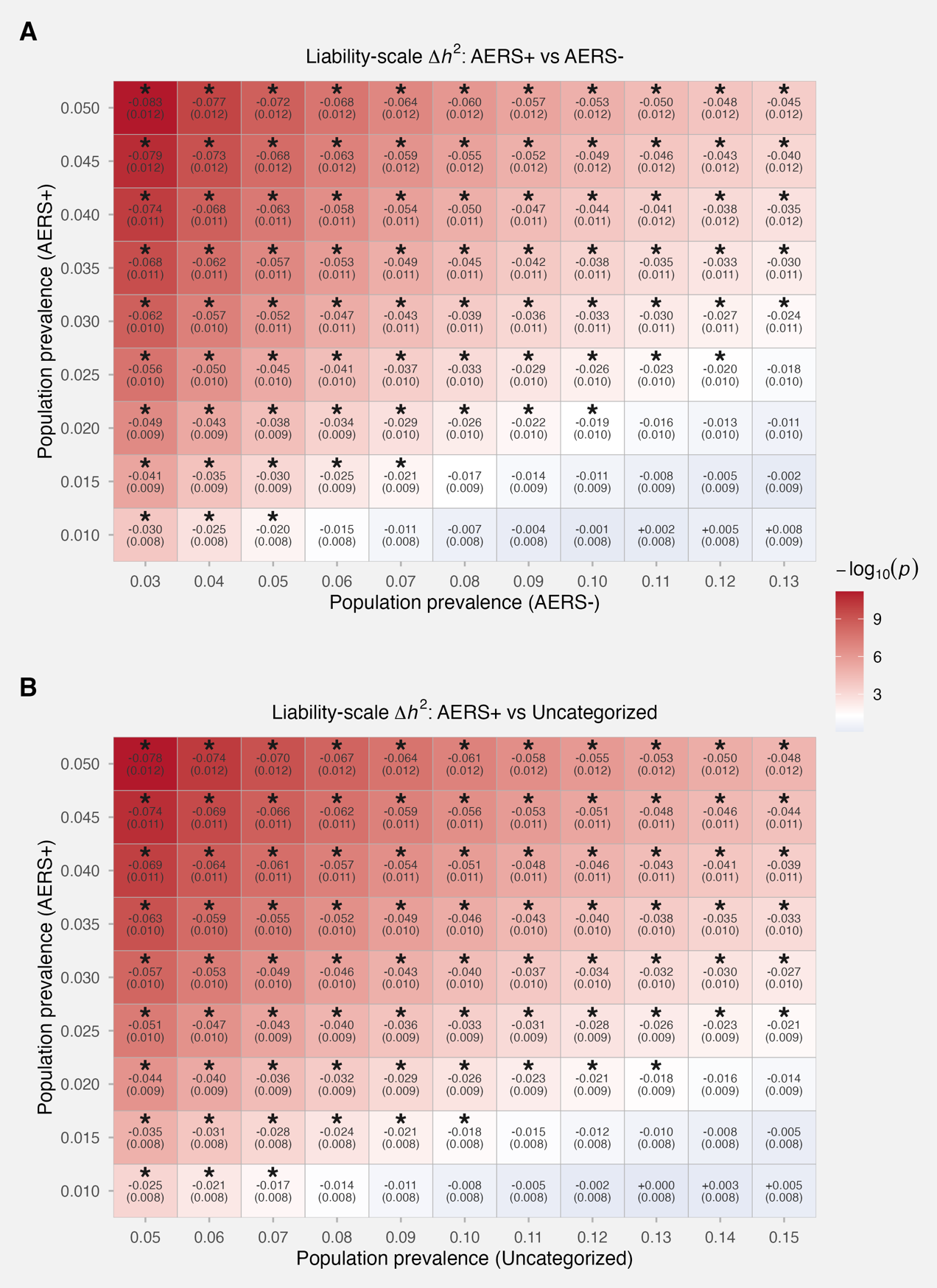

**Figure 4:** Liability-scale SNP-heritability differences between AERS subtypes across a grid of plausible population prevalences, estimated with a block-jackknife. Each cell reports the difference in liability-scale h² (Δh²) with its jackknife standard error in parentheses, and is coloured by -log10(p) for the jackknife test that Δh² = 0; white corresponds to p = 0.05. (A) AERS+ vs Uncategorized; (B) AERS+ vs AERS-. The y-axis sweeps AERS+ population prevalence; the x-axis sweeps the comparator subtype's prevalence.

##### Supplementary Figure 5

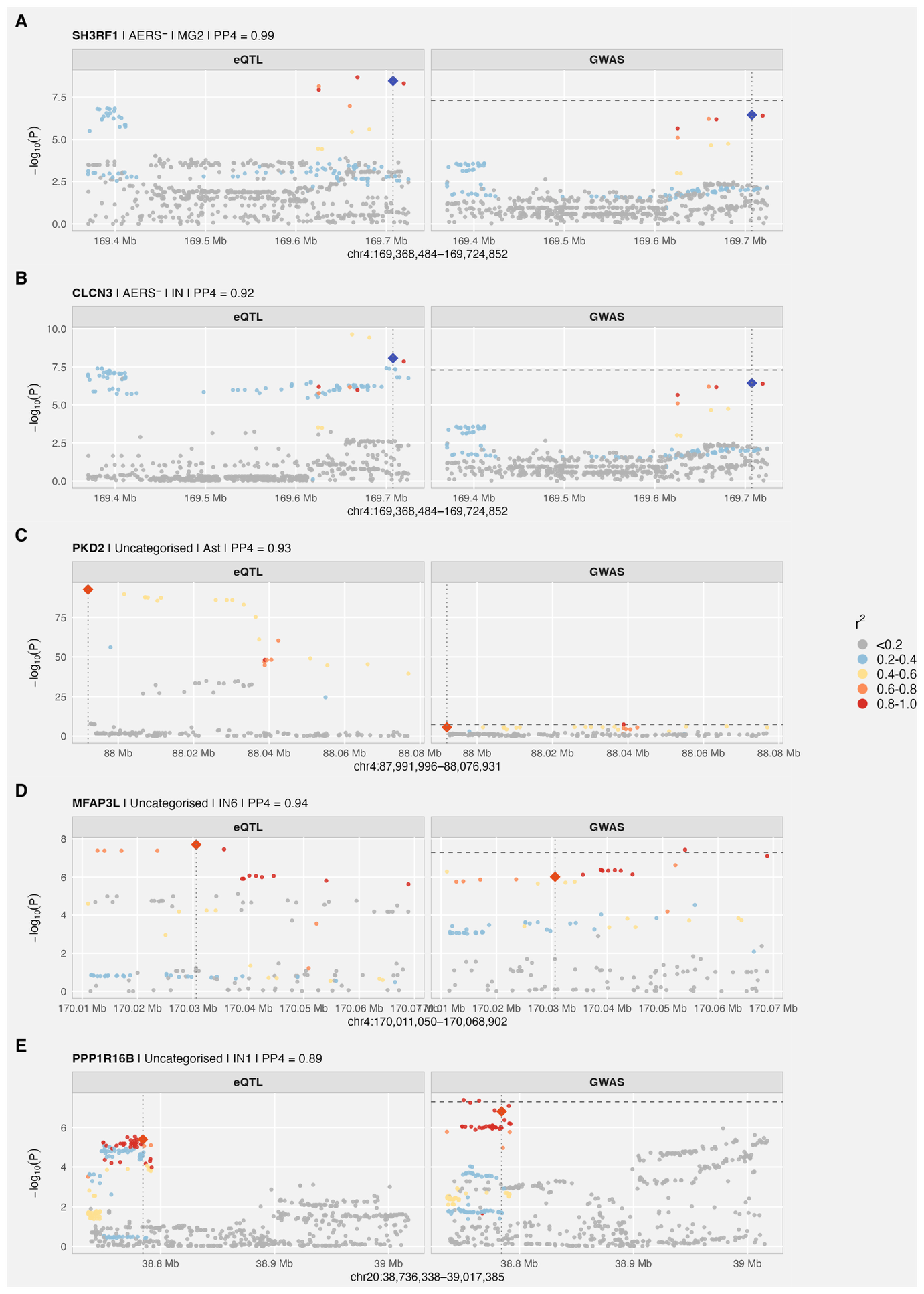

**Figure 5:** Regional colocalization plots for every eQTL colocalization, beyond the three loci shown in main Figure 2. Each panel shows the GWAS signal (top) and the matched eQTL signal (bottom); points are coloured by LD (r²) to the lead GWAS variant, which is marked as a filled diamond. The dashed horizontal line marks P = 5 × 10⁻⁸. Panel titles list the colocalised gene, AERS subtype, eQTL tissue or cell type, and the coloc posterior probability of a shared causal variant (PP.H4).

##### Supplementary Figure 6

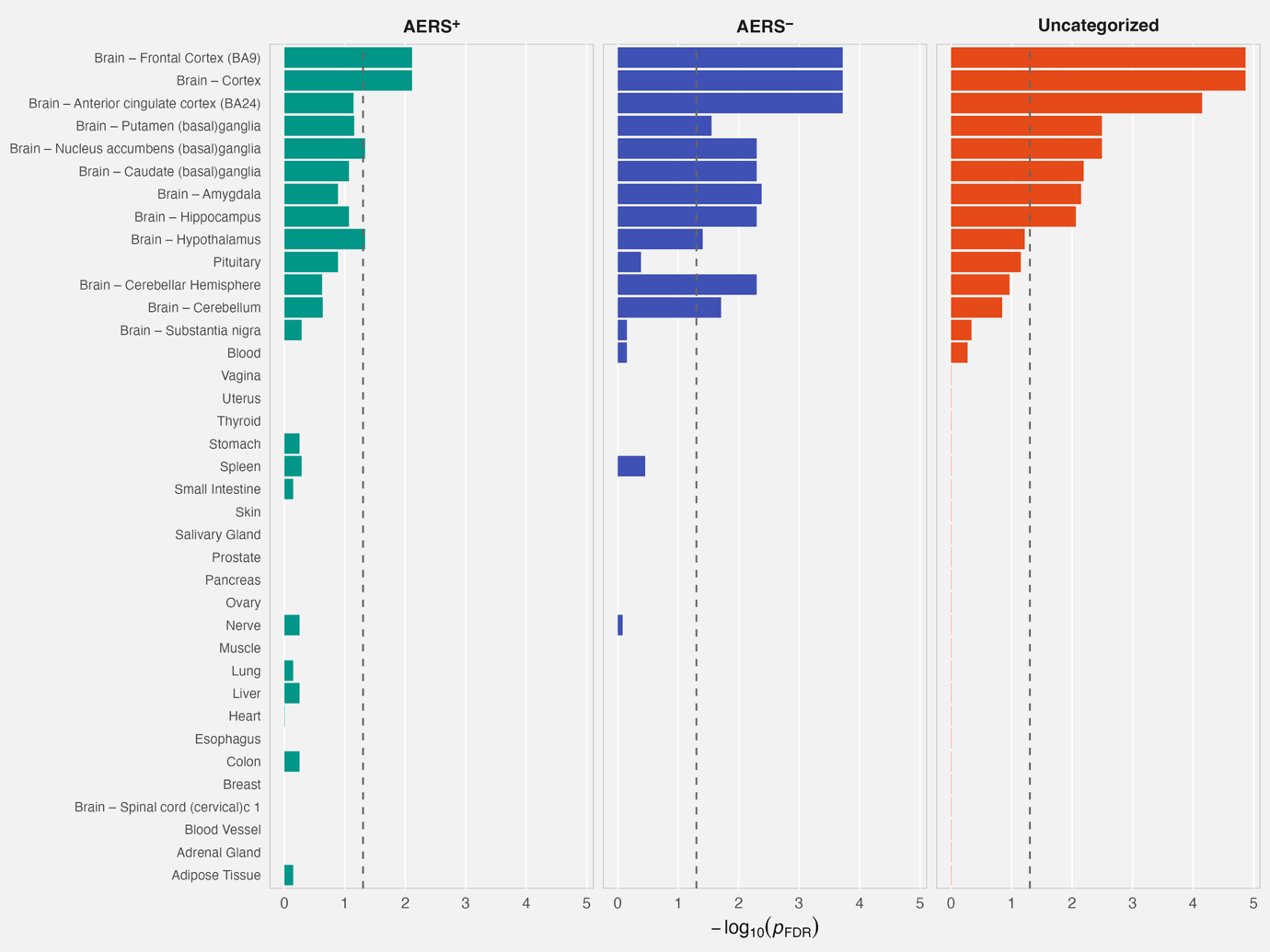

**Figure 6:** Stratified-LDSC partitioned heritability across 37 GTEx tissues for AERS+, AERS-, and Uncategorized MDD. Bars show -log10 of the FDR-adjusted enrichment p-value (FDR applied within phenotype across the 37 tissues). The dashed line marks p_FDR = 0.05.

##### Supplementary Figure 7

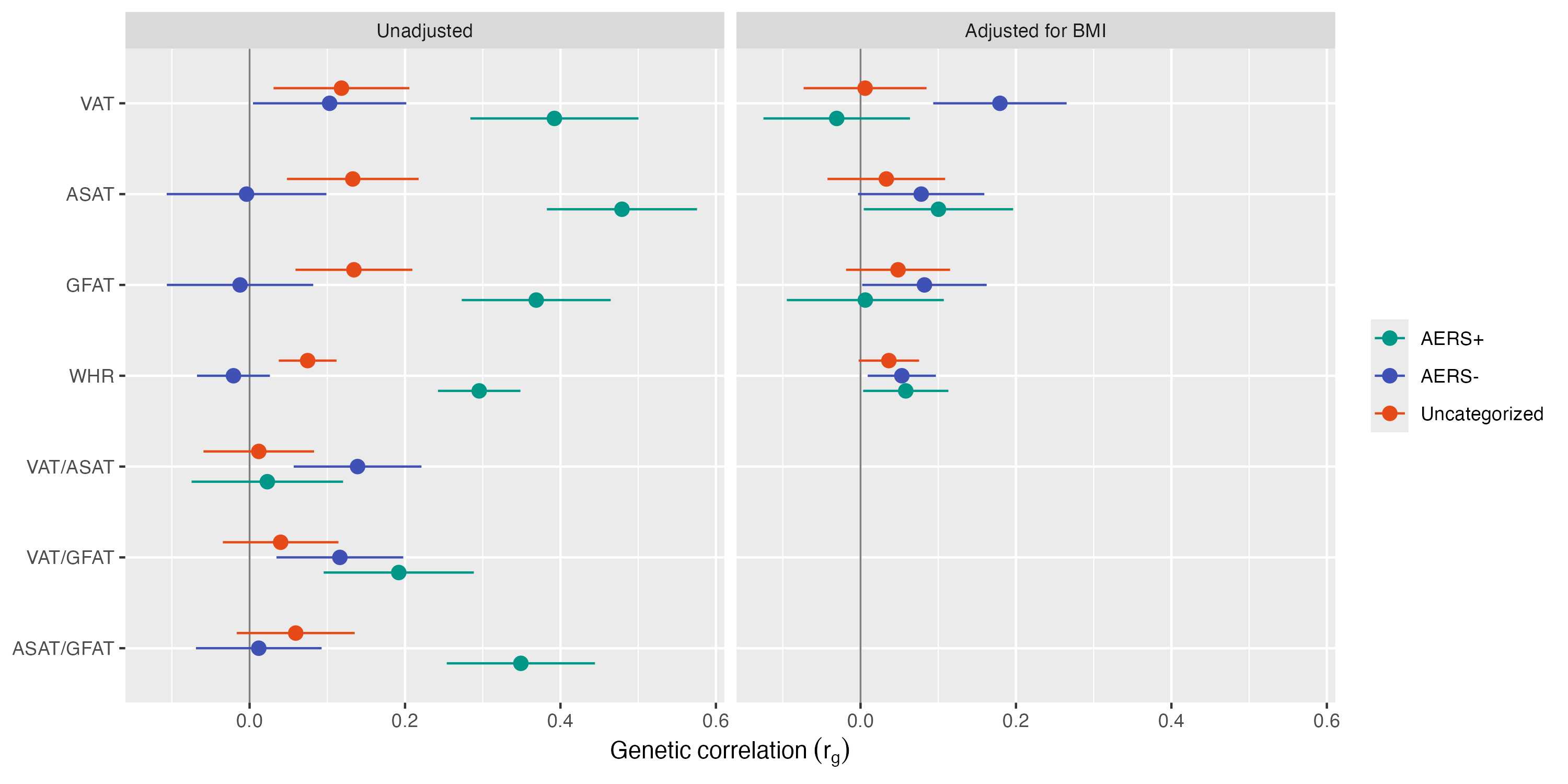
**Figure 7:** Genetic correlations of AERS+, AERS-, and Uncategorized MDD with abdominal MRI-derived fat depots (visceral, VAT; abdominal subcutaneous, ASAT; gluteofemoral, GFAT), their pairwise ratios, and anthropometric waist-to-hip ratio (WHR), shown side-by-side for unadjusted GWAS and depot/WHR GWAS adjusted for BMI. The BMI-adjusted panel only shows VAT, ASAT, GFAT, and WHR because BMI-adjusted GWAS are not available for the ratio traits. Error bars are 95% confidence intervals; the grey vertical line marks rg = 0.

##### Supplementary Figure 8

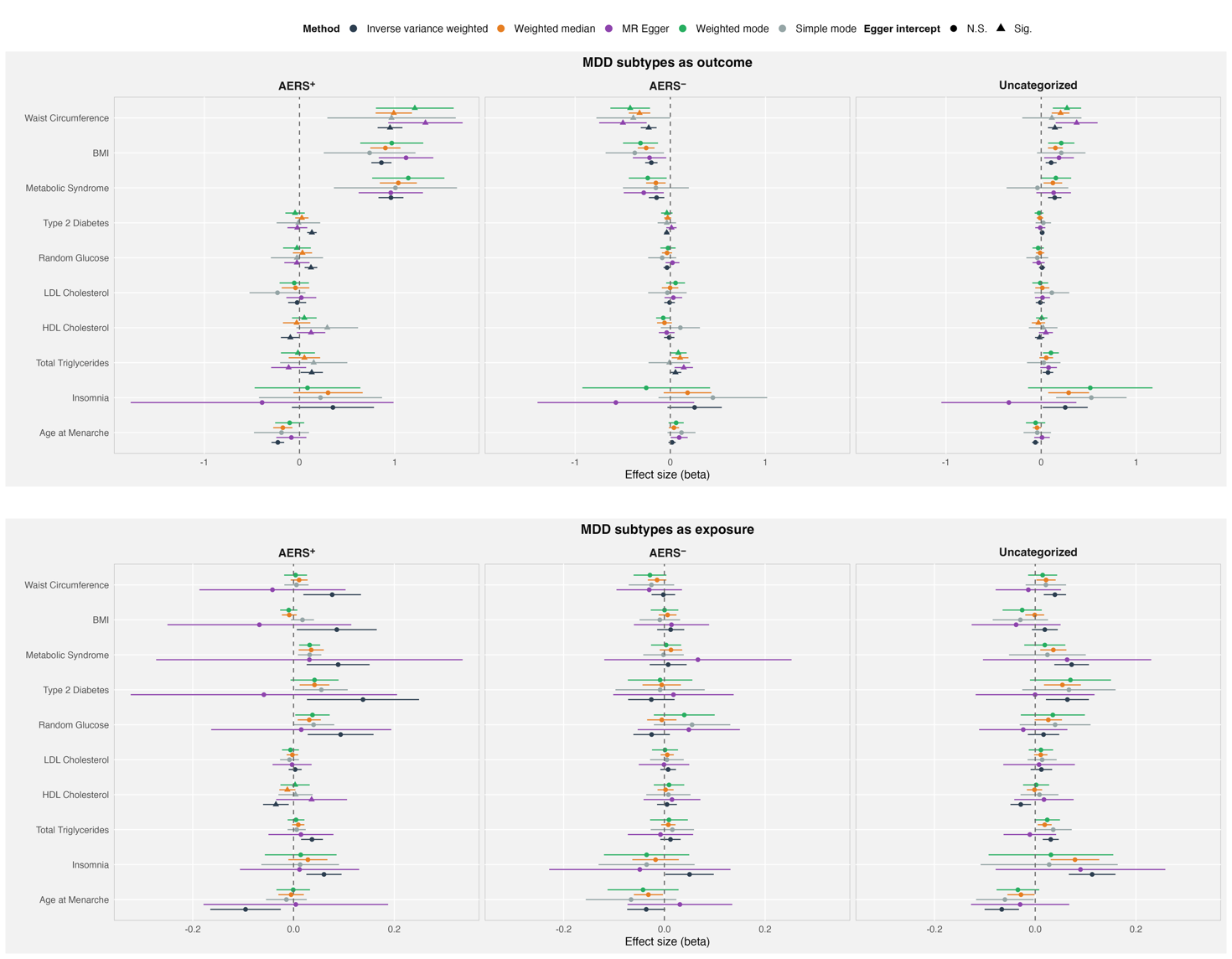

**Figure 8:** Two-sample Mendelian randomization results for AERS+, AERS-, and Uncategorized MDD in both directions (subtype as exposure and as outcome). Effect estimates are shown for five estimators (inverse-variance weighted, MR-Egger, weighted median, weighted mode, simple mode). Error bars are 95% confidence intervals; the dashed line marks β = 0.

### Cohorts

#### GLAD+ and UK Biobank

##### Cohort description

The NIHR BioResource Genetic Links to Anxiety and Depression (GLAD) Study is an online study of adults living in the UK with lifetime experience of depression and/or anxiety^7^. Participants have completed extensive online questionnaires on sociodemographics, mental and physical health, and lifestyle. Currently, the GLAD study has over 66,000 consented participants, with more than 50,000 having completed the online survey, and over 36,000 saliva samples collected. During the COVID-19 pandemic, the GLAD Study research team recontacted GLAD participants and healthy volunteers from other NIHR BioResource (<https://bioresource.nihr.ac.uk/>) studies to conduct the Covid-19 Psychiatry and Neurological Genetics (COPING) study, including >20,000 participants with psychiatric disorders and >11,000 healthy volunteers, two-thirds of whom have been genotyped. “GLAD+” consists of GLAD as well as participants from the COPING study, who completed the same measures but were not recruited on the basis of mental health diagnostic status. UK Biobank (UKB) is a UK population-based health study^8^. Eligible participants were aged between 40 and 69 at recruitment and answered touchscreen, online and verbal questions, as well as agreeing to medical record linkage for general practitioner (GP) READ v2/v3 codes and hospital inpatient ICD-10 codes. A subset of participants completed one or both of two follow-up online mental health questionnaires (MHQ) in 2016 and 2022^9,10^. More than 206,000 UK Biobank participants have data for either MHQ1 or MHQ2. Both MHQ1 and MHQ2 measured lifetime depression, anxiety disorders, and other psychiatric disorders.

##### Phenotype definition

###### GLAD+ phenotype definition

In GLAD+, DSM Criterion A symptom burden was operationalised using eight CIDI-derived depressive symptom indicators: depressed mood, anhedonia, weight/appetite change, sleep change, fatigue, feelings of worthlessness, concentration difficulties, and thoughts of death. MDD was defined as ≥5 of these symptoms, together with functional impairment, symptoms lasting most or all of the day, and symptoms occurring almost every day or every day. AERS+ cases were defined by co-occurring weight/appetite gain and hypersomnia, whereas AERS− cases were defined by co-occurring weight/appetite loss and hyposomnia. Remaining MDD cases were classified as uncategorised. Controls were selected from COPING_NBR participants without CIDI-derived MDD. Analyses were restricted to genotyped individuals. Participants with psychotic disorders, bipolar disorder, schizophrenia, or personality disorders were excluded from both cases and controls, and participants who self-reported lifetime MDD episodes or anxiety disorders were additionally excluded from the control group.

###### UKBB phenotype definition

Within the larger UK biobank cohort we analyzed all individuals that participated either in the first or second instance of the mental health questionnaire (“MHQ1” or “MHQ2”). We further restricted our analysis to individuals with genotype data, reported sex equal to genotyped sex, of European ancestry and a sample missing rate of less than 2%. We further excluded from analysis individuals with any ICD code for schizophrenia or bipolar disorder (F20-F29, F30, F31) or who have been prescribed lithium or antipsychotics (field 20003). We defined controls based on further criteria: 0 reported depressive symptoms across MHQ1 and MHQ2 (fields 20441, 20446,29011, 29012), no self-report of MDD (fields 20002, 20544, 29000), no ICD code for depression (field 41270, F32-F34, F38, F39) and not classified as probable depression from the baseline measurements (field 20126). We defined lifetime MDD within each instance of the MHQ based on the presence of >= 5 DSM-V symptoms with frequency of depressed days: “almost every day” or “every day”, Fraction of day depressed: “Most of the day” or “All day long” and Impact on life; “somewhat” or “a lot” . We incorporated the repeat measurements across MHQ1 and MHQ2 in the following manner:

Within each instance of the MHQ, we defined AERS +, AERS - and Uncategorised.

We considered the union of individuals that qualified as AERS + in either MHQ1 or MHQ2 as AERS (+). From this union set we removed individuals who met criteria for AERS – in the second measurement (for example, AERS + in MHQ1 but AERS (-) in MHQ2, or vice versa), resulting in the final set of individuals meeting criteria for AERS (+)

We subsequently defined AERS (-), and removed any individuals that had already qualified as AERS (+). Lastly, we defined Uncategorised as any individuals meeting criteria for MDD, but who had not been classified as AERS (+) or AERS (-).

All data from GLAD+ study were genotyped by ThermoFisher on the UK Biobank Axiom Array v1 and v2 across numerous genotyping batches. Genetic data were restricted to individuals from European ancestries (749,044 SNPs before quality control). Ancestry was determined using GenoPred^11^, by projecting GLAD+ individuals on genomic principal components from the 1000 Genomes reference data, and assigning individuals a genetic ancestry if they lay < 3SD from the mean of individuals from that ancestry superpopulation. Quality control exclusions were conducted for variants with MAF < 0.01, call rate < 0.95, or deviant from Hardy-Weinberg equilibrium (p < 1×10⁻⁸). Individuals were excluded if they: had withdrawn from the study following genotyping, were a duplicate of a higher-quality sample (not including known identical twins), were known to be mislabelled, their genotypic sex (males Fx > 0.8, females Fx < 0.5) did not match their sex assigned at birth, were outliers on genome-wide heterozygosity (|F-hat| > 0.2), or had an excess of relatives (average pi-hat > 3SD from the mean). Following quality control, 33,635 individuals and 484,182 variants were available for imputation. Imputation was carried out to TopMED Freeze 8^12^. Data was then further restricted to MAF ≥0.01 and R2 ≥ 0.3, leaving 15,009,228 variants for analysis.

UKB participants provided blood samples, genotyped on the UK Biobank Axiom Array (90% of the sample) or the UK BiLEVE array. Several sample exclusions were implemented based on quality control procedures performed by the UKB and described elsewhere (i.e., missing rate, heterozygosity, sex discordance)^8^. In addition, we excluded participants with genotype missingness > 0.02 and related individuals identified as KING > 0.04, excluding one participant from each related pair, while preferentially selecting from each pair those participants that answered the MHQ. Imputed dosage data for 487,422 participants based on 670,739 markers was provided by the UKB. Genetic variants were imputed using IMPUTE4 software with the Haplotype Reference Consortium reference panel^13^, 1000 Genomes phase 3^14^, and UK10K Consortium reference panel^15^. SNP quality control involved the exclusion of imputed SNPs with MAF < 0.01 and INFO < 0.4. After quality control procedures we obtained a sample size of 413,259 participants of mixed ancestry and 9,830,370 imputed genotypes. Ancestry classification into super populations was implemented using 4 means-clustering on the first two ancestry-informed principal components, identifying 397,319 European ancestry participants.

To maximise power in this case-control analysis, we pooled the participants into a single sample, with GLAD+ providing most cases, and UKB the controls. We merged genotype data from GLAD+ and UKB using Plink 1.9. For the GWAS analysis, we performed a final round of quality control on the combined sample, restricting to variants with MAF > 0.01, missing rate < 0.02, Hardy-Weinberg equilibrium exact test p-value > 1×10⁻⁸, and individuals with missingness < 0.02.

We used KING to assess relatedness between GLAD+ and UKB participants. A total of 1,815 participant pairs shared identical genetic data; we assumed that pairs within the same dataset represent twins, while pairs across the two datasets represent duplicates (n = 1,633). Where a participant had provided data for both studies (‘duplicates’), we preferentially retained the higher-quality UKB blood-based genotype data, complete phenotype data, and evidence of meeting case criteria, and removed the other data for that participant. Principal components were projected onto the whole pooled sample from unrelated individuals using flashpca^16^.

Ethical approval for the GLAD Study and NBR COPING study was obtained from the London-Fulham Research Ethics Committee (REC reference 18/LO/1218 and 20/SW/0078, respectively, COPING project no. 282754.). The dataset used in the present analysis was freeze 2023-06-07. UKB analyses were conducted under application ID 82087 and 22224. All participants provided informed consent.

##### Statistical analysis

The GWAS was performed in REGENIE^17^ version 3.1.3 with covariates of the batch and the first 10 genetic principal components.

#### BIONIC

##### Cohort description

The BIObanks Netherlands Internet Collaboration (BIONIC) is a nationwide project uniting

16 Dutch population-based and clinical cohorts to investigate the genetics of lifetime major

depressive disorder (MDD)^18^. The collaboration was established within the Biobanking and

BioMolecular resources Research Infrastructure (BBMRI-NL) to standardize phenotyping

and integrate genetic data across existing Dutch biobanks. The participating cohorts, in

alphabetical order, are: the Doetinchem Cohort Study^19^, the Hoorn Study, the New Hoorn

Study, and the Hoorn Diabetes Care System cohort^20^, Longitudinal Aging Study

Amsterdam^21^, Lifelines ^22^, MOod Treatment with Antidepressants or Running ^23^,

MooDFOOD ^24,25^, Nijmegen BIomedische Studie ^26^, Netherlands Study of Depression and

Anxiety and Netherlands Study of Depression and Anxiety sibling cohort ^27^, Netherlands

Study of Depression in Older Persons ^28^, Nutrition Questionnaires plus ^29^, Netherlands Twin

Register ^30^, Tracking Adolescents’ Individual Lives Survey and Tracking Adolescents’

Individual Lives Survey clinical cohort ^31^. Phenotyping of MDD was primarily conducted using the Lifetime Depression Assessment Survey (LIDAS), an online DSM-5–based instrument validated against the Composite International Diagnostic Interview (CIDI). Lifetime MDD status was determined for 123,950 participants (29,650 cases and 94,300 screened controls), of whom 64,941 individuals of European ancestry had genome-wide SNP data available. Genotyping was performed on multiple Illumina and Affymetrix arrays and harmonized via a centralized quality-control and imputation pipeline at the high-performance computing server of the Genomics Coordination Center (Groningen). Samples with call rate <90%, sex discordance, excess heterozygosity, or relatedness were excluded; variants were filtered for call rate <95%, HWE p<1×10⁻⁴, and MAF <1%. Data were aligned to genome build HG19/GRCh37 and imputed to the Haplotype Reference Consortium (v1.1) using BEAGLE 5.1.

##### Phenotype definition

Lifetime MDD data were derived from clinical interviews and depression self-report questionnaires, including the LIDAS, the Mini International Neuropsychiatric Interview (MINI), the CIDI, and the Diagnostic Interview Schedule (DIS) for identification of cases and controls, and the Adult Self-Report of the Achenbach System of Empirically Based Assessment (ASR-ASEBA), the Beck Depression Inventory (BDI), the Hospital Anxiety and Depression Scale (HADS), and the Center for Epidemiologic Studies Depression Scale (CES-D) for a subset of controls. MDD cases were defined in accordance with DSM-5 criteria as ever having had a ≥2-week period with significant dysfunction where five out of nine depression symptoms were present, including at least one core symptom. MDD controls were defined in the absence of such a period, or having a normalized T-score sumscore lower than 60 on the self-report questionnaires (and no counterindication on any other instrument). Controls were additionally screened for the diagnosis or treatment of psychopathology other than depression when such information was available, including generalized anxiety disorder, bipolar disorder, attention-deficit (hyperactivity) disorder (AD(H)D), alcohol addiction, drug addiction, personality disorder, and antidepressant use. AERS subtypes were defined based on weight/appetite gain and hypersomnia and insomnia.

##### Statistical analysis

Genome-wide association studies were performed using GCTA fastGWA (v1.94.1), adjusting for biological sex, age at measurement, six genotype array dummies to account for the seven genotype arrays used across cohorts, and the first 10 ancestry-informative principal components projected from the 1000 Genomes Project (Phase 3 v5) reference panel. Relatedness in the sample was addressed with a sparse genetic relationship matrix.

#### Estonia Biobank

##### Cohort description

The Estonian Biobank (EstBB) is a volunteer-based cohort (N=212,000) comprising omics and extensive health data from national electronic health records for each participant. Diagnostic information (ICD-10 codes) and prescription records (ATC codes, prescription status, and purchase dates) are available for all individuals through linkages to electronic health records, including the National Health Insurance Fund’s database^32^. In 2021, the Mental Health online Survey (MHoS) was administered to collect lifestyle and mental health symptom data from 86,000 participants ^33^. Genotyping was performed at the Core Genotyping Lab, Institute of Genomics, University of Tartu, using Illumina Global Screening Arrays (GSAv1.0, GSAv2.0, and GSAv2.0_EST). Individuals with a call rate below 95% or sex discrepancies between genotype and phenotype data were excluded. Variant-level quality control removed markers with call rate below 95% or Hardy–Weinberg equilibrium p-value < 1×10⁻⁴ (autosomal variants only). Imputation was conducted using an Estonian-specific reference panel based on 2,695 whole-genome sequenced individuals.

The activities of the EstBB are regulated by the Human Genes Research Act, which was adopted in 2000 specifically for the operations of the EstBB. Individual level data analysis in the EstBB was carried out under ethical approval [1.1-12/4561] from the Estonian Committee on Bioethics and Human Research (Estonian Ministry of Social Affairs), using data according to release application [P20 6-7/GI/24461] from the Estonian Biobank.

##### Phenotype definition

We analyzed all individuals that participated in the MHoS. We further excluded from analysis individuals with any ICD code for schizophrenia or bipolar disorder (F20-F29, F30, F31) or who have been prescribed lithium or antipsychotics. We defined controls based on further criteria: 0 reported depressive symptoms across MHoS, and no ICD code for depression (F32-F33). We defined lifetime MDD within each instance of the MHoS based on the presence of >= 5 DSM-V symptoms with frequency of depressed days: “almost every day” or “every day” and an impact on life; “somewhat” or “a lot”. We classified individuals reporting both hypersomnia and weight gain as AERS+, individuals reporting insomnia and weight loss as AERS-, and all remaining individuals meeting criteria for MDD as Uncategorised.

##### Statistical analysis

Genome-wide association studies were conducted using REGENIE v 4.0, adjusting for the first 10 principal components, age and biological sex.

#### Australian genetics of depression study (AGDS)

##### Cohort description

The Australian Genetics of Depression Study (AGDS) includes participants with self-reported diagnoses of depression, recruited via two complementary approaches: a national media campaign inviting volunteers with a history of depression, and identification through Australian government prescription records^34^. Participants completed online questionnaires within several modules, which consisted of a core module (primarily assessing depression, essential information on self-report mental health diagnoses, medication response and side effects) and 10 ten additional ‘satellite’ modules that assessed a range of complex traits of relevance to mental health using a variety of scales and questionnaires. Control participants were selected from the QSkin Sun and Health Study (QSkin) conducted at the QIMR Berghofer Medical Research Institute. QSkin is a prospective cohort study initiated in 2011 primarily to examine skin cancer outcomes. Participants aged 40 to 70 years responded to a mailing to residents of Queensland, Australia, selected at random from the electoral role (n = 43 794). A total of 17 218 QSkin participants provided a saliva sample in 2014; answered the lifestyle questionnaire, which included a disease checklist comprising questions about ever having been diagnosed with psychiatric disorders; and provided consent for their data to be used for future research. All protocols and questionnaires for both the AGDS and QSkin cohorts were approved by the QIMR Berghofer Medical Research Institute Human Research Ethics Committee (P2118, P1309 and P2034).

Genotyping was conducted using the Illumina Infinium Global Screening Array platform. Prior to imputation, a common set of high QC markers between the different genotyping batches was obtained. Marker exclusion criteria included: unknown or ambiguous map position and strand alignment in a BLAST search, missingness >5%, p(HWE test)< 10^-6), MAF<1%, GenTrain score <0.6. The Michigan imputation server was used to impute the genotypes using the HRCr1.1 as a reference panel. Individuals were excluded based on a high missingness (missing rate > 3%), inconsistent (and unresolvable) sex, or if deemed ancestry outliers from the European population (6 standard deviations from the first two genetic principal components from 1000 Genomes).

##### Phenotype definition

MDD case status was based on meeting DSM criteria (presence of ≥5 symptoms, together with functional impairment, symptoms lasting most or all of the day, and symptoms occurring almost every day or every day), assessed with the CIDI-SF (Composite International Diagnostic Interview - Short Form). AERS+ cases were defined as MDD cases who reported both weight/appetite gain and hypersomnia. AERS− cases were defined as MDD cases who reported both weight/appetite loss and hyposomnia. Remaining MDD cases were classified as uncategorised. Controls were selected as QSkin participants who reported not having been diagnosed, experienced or treated for depression nor experienced postnatal or antenatal depression. Participants were excluded if they reported a psychiatric comorbidity (self-reported diagnosis of schizophrenia, bipolar disorder, a personality disorder, or a substance use disorder), were of non-European ancestry, or had a relative in the study (removed 1 individual per pair with pi-hat > 0.2).

##### Statistical analysis

GWAS analysis of the three phenotypes were conducted using a logistic regression (--glm) in PLINK2 (v2.00a3.3LM) using the default settings. Imputed genotype dosages for variants with minor allele frequency > 0.5% were used for the analyses. The following covariates were included in the analyses: age, sex, and the first 10 ancestral principal components.

#### The Swedish Twin Registry (YATSS and STAGE)

##### Cohort description

The Swedish Twin Registry is a nation-wide population-based registry that includes twins born from 1886 and onwards. The registry combines questionnaire/interview data with DNA , and other biological samples, and data linkage to the national health registers, making it a valuable platform for genomic research. Detailed information about STR are described elsewhere^35–37^. Briefly, two subcohorts, i.e. the Study of Twin Adults: Genes and Environment (STAGE) and the Young Adult Twins in Sweden Study (YATSS) were included in the AERS GWAS meta-analysis. STAGE was designed to assess common complex diseases and relevant environmental exposures in young and middle adulthood and invited approximately 42,582 twins born 1958-1985 to participate in web-based questionnaires or telephone interviews during 2005-2006 with an overall response rate close to 60%. YATSS was designed to collect both early-life exposures and common health problems in young adulthood, and invited 16,244 twins born 1986-1992 to participate online or by telephone in 2013 with response rate of 42%. The ethical approval has been granted by the Swedish Ethical Review Authority (Dnr 2023-03073-01). DNA extracted from saliva samples of STAGE and YATSS participants was genotyped on Illumina arrays (Global Screening Array Multi-Disease BeadChip) at the SNP&SEQ Technology Platform (Uppsala, Sweden). Genotype data were then processed with standard Ricopili pipeline for quality control: excluding samples with low call rate <0.98, excess heterozygosity with FHET outside ±0.2 , sex mismatch, and variants with low call rate, invariant, Hardy–Weinberg disequilibrium (P < 1 × 10^−6^ in controls and P < 1 × 10^−10^ in cases), or differential missingness between cases and controls. After QC, genotyped data were then imputed to the Haplotype Reference Consortium reference panel (release 1.1). Genotypes for monozygotic cotwins were inferred from their genotyped twin.

##### Phenotype definition

In YATSS and STAGE, lifetime MDD was defined based on 5 >= out of the nine DSM-V symptoms, with at least 1 cardinal symptom endorsed and moderate to large impact on daily life. Hypersomnia, hyposomnia, weight gain and weight loss were used to classify lifetime MDD individuals into AERS+, AERS- and Uncategorised. Controls were required to have 0 reported depressive symptoms.

##### Statistical analysis

GWAS were performed in the YATSS and STAGE genotyped samples using the REGENIE^17^. Models were adjusted for standardized birth year, sex, and the first 10 genetic principal components. For step 1, we used variants filtered at MAF ≥ 1%, MAC ≥ 200, and Hardy–Weinberg equilibrium P ≥ 1 × 10^-15^ to generate genetic relatedness matrices; For step 2, no SNP-level filtering was applied to the imputed data at the GWAS stage beyond the upstream QC and post-imputation filtering described above.

#### Generation Scotland

##### Cohort description

The cohort is described in detail in ^38^. Generation Scotland; Scottish Family Health Study (GS:SFHS) includes approximately 24,000 adults (aged 18 and older) from 7,000 families recruited from the general Scottish population between 2006 and 2011. Around 90% of participants attended assessment clinics in Dundee, Glasgow, or Aberdeen for biological sampling and clinical testing, while the remainder provided questionnaire data and saliva samples by mail. The study’s primary goal is to explore the genetic and environmental determinants of common diseases, with a particular emphasis on mental health. Ethical approval for the study was obtained from the Scotland A Research Ethics Committee (REC reference number 14/55/0039) and the local Research and Development offices. All participants provided written informed consent prior to the collection of any data or samples.

##### Phenotype definition

As part of STratifying Resilience and Depression Longitudinally (STRADL) participants in GS:SFHS were recontacted and completed the CIDI-SF questionnaire (N=9,618)^39^. Lifetime MDD was diagnosed using the Structured Clinical Interview for DSM Disorders (endorsing either question 1 and/or 2, and four or more additional symptoms). Individuals with a CIDI-SF diagnosis of bipolar disorder or hypomania were excluded. Subgroup membership was defined using the CIDI questions relating to weight (gain and loss) and sleep change (hyposomnia was defined from questions related to trouble sleeping and waking early) during their worst depressive episode. No data was available on appetite change. Individuals with diagnoses of lifetime MDD but that did not meet criteria for AERS+ or AERS- were defined as uncategorised.

##### Statistical analysis

An unrelated subset of STRADl was defined using KING-robust kinship estimator to remove individuals third degree or closer (--king-table-filter 0.05). The association analysis was performed using HRC v1.1 imputed genotype data after removal of monogenic and SNPs with INFO scores < 0.8 and PLINK 1.9 fitting age, sex and six genetic principal components under an additive model^40^.

#### Psychiatric Genomics Consortium (PGC)

##### Cohort description

Data was drawn from the following cohorts: QIMR (qi3c, qi6c, qio2), RADIANT (rad3), STAR*D (stm2), GenRED (grnd). Symptoms were assessed by trained interviewers using structured diagnostic instruments and DSM checklists. Information on cohort genotyping and other processing steps have been described in detail in a previous article^41^. The genotypes were processed through Ricopili^42^ (with the following QC: SNP missingness < 0.05; sample missingness < 0.02; autosomal heterozygosity deviation (|Fhet|<0.2); and SNP Hardy-Weinberg equilibrium (P>10^−6^ in controls, P>10^−10^ in cases). QC'd genotypes were then imputed to the 1000 Genomes Reference Panel. Cohorts "grnd", "stm2", "rad3", "qi3c", "qi6c", "qio2" were selected on presence of both weight loss/increase and sleep increase/decrease, without overlap in other cohorts. Cohorts qi3c, qi6c, qio2 were excluded in the analysis of AERS+ due to insufficient number of cases for GWAS.

##### Phenotype definition

MDD was determined based on cohort specific sources. Additional phenotype information on weight decrease/increase and sleep increase/decrease was used to group MDD individuals into AERS+ (increased weight & increased sleep), AERS- (decreased weight and decreased sleep). Individuals not meeting criteria for AERS+ or AERS- were classified as Uncategorised.

##### Statistical analysis

We used plink2 with arguments --glm, --mac 30 to perform logistic regression, with the first five principal components as covariates. We subsequently filtered on MAF >= 10% as we observed substantial inflation in test statistics for lower frequency variants. All PGC cohorts were subsequently meta-analysed with tidyGWAS, and only variants with effective sample size >= 0.6*max(EffectiveN) were retained.

#### Twins early development study (TEDS)

##### Cohort description

The Twins Early Development Study (TEDS) is a longitudinal twin study that recruited over 16,000 twin-pairs born between 1994 and 1996 in England and Wales through national birth records^43^. More than 10,000 of these families are still engaged in the study. TEDS was and still is a representative sample of the population in England and Wales. Rich cognitive and emotional/behavioural data have been collected from the twins from infancy to emerging adulthood, with data collection at first contact and at ages 2, 3, 4, 7, 8, 9, 10, 12, 14, 16, 18 and 21. Samples were removed from subsequent analyses on the basis of call rate (<0.98), suspected non-European ancestry, heterozygosity, and relatedness other than dizygotic twin status. SNPs were excluded if the minor allele frequency was smaller than 0.5%, if more than 2% of genotype data were missing, or if the Hardy Weinberg p-value was lower than 10^-5^. Non-autosomal markers and indels were removed. Association between SNP and the platform, batch, plate or well on which samples were genotyped was calculated; SNPs with an effect p-value < 10^-4^ were excluded. A total sample of 10,346 samples (including 3,320 dizygotic twin pairs and 7,026 unrelated individuals), with 7,289 individuals and 559,772 SNPs genotyped on Illumina and 3,057 individuals and 635,269 SNPs genotyped on Affymetrix remained after quality control. Samples were removed from subsequent analyses on the basis of call rate (<0.98), suspected non-European ancestry, heterozygosity, and relatedness other than dizygotic twin status. SNPs were excluded if the minor allele frequency was smaller than 0.5%, if more than 2% of genotype data were missing, or if the Hardy Weinberg p-value was lower than 10-5. Non-autosomal markers and indels were removed. Association between SNP and the platform, batch, plate or well on which samples were genotyped was calculated; SNPs with an effect p-value < 10^-4^ were excluded. A total sample of 10,346 samples (including 3,320 dizygotic twin pairs and 7,026 unrelated individuals), with 7,289 individuals and 559,772 SNPs genotyped on Illumina and 3,057 individuals and 635,269 SNPs genotyped on Affymetrix remained after quality control. Imputation: Genotypes from the two platforms were separately phased using EAGLE2 and imputed into the Haplotype Reference Consortium (release 1.1) using the Positional Burrows-Wheeler Transform method through the Sanger Imputation Service. Prior to merging, we excluded variants with info <0.75 and removed non-overlapping SNPs between platforms. After merging, we tested for minor allele frequency differences between platforms and removed SNPs with an effect p-value < 10⁻⁴, and Hardy Weinberg p-value > 10⁻⁵.

TEDS is supported by the UK Medical Research Council (MR/V012878/1 and previously MR/M021475/1). Ethical approval for TEDS is provided by the Ethics Committee (reference: PNM/09/10–104). Written informed consent was obtained prior to each wave of data collection from parents and from twins themselves from age 16 onwards.

##### Phenotype definition

Lifetime MDD was defined using the CIDI-SF questionnaire. To meet criteria, individuals had to report 5/9 DSM-V symptoms, with symptoms present most of the day, most of the week and a moderate to large impact on roles in daily life. Controls were required to have 0 self-reported lifetime depressive symptoms. Among individuals meeting MDD criteria, individuals reporting both hypersomnia and weight gain were classified as AERS+, individuals reporting insomnia and weight loss as AERS-, and all remaining individuals as Uncategorised.

##### Statistical analysis

GWAS was performed with REGENIE, using the first 10 principal components as covariates to adjust for population stratification.

### Major Depressive Disorder Working Group of the Psychiatric Genomics Consortium

Mark J Adams 1

Fabian Streit 2, 3, 4, 5

Xiangrui Meng 6

Swapnil Awasthi 7

Brett N Adey 8

Karmel W Choi 9, 10

V Kartik Chundru 11, 12

Jonathan RI Coleman 8, 13

Bart Ferwerda 14

Jerome C Foo 2, 15, 16, 17

Zachary F Gerring 18

Olga Giannakopoulou 6

Priya Gupta 19, 20

Alisha S M Hall 2, 21

Arvid Harder 22

David M Howard 8

Christopher Hübel 8, 23, 24

Alex S F Kwong 1, 25

Daniel F Levey 19, 20

Brittany L Mitchell 18, 26, 27, 28

Guiyan Ni 29

Vanessa K Ota 30

Oliver Pain 31

Gita A Pathak 19, 32

Eva C Schulte 33, 34, 35, 36, 37

Xueyi Shen 1

Jackson G Thorp 18

Alicia Walker 29

Shuyang Yao 22

Jian Zeng 29

Johan Zvrskovec 8, 13

Dag Aarsland 38

Ky'Era V Actkins 39

Mazda Adli 40, 41

Esben Agerbo 24, 42, 43

Mareike Aichholzer 44

Allison Aiello 45

Tracy M Air 46

Thomas D Als 43, 47, 48

Evelyn Andersson 49

Till F M Andlauer 50, 51

Volker Arolt 52

Helga Ask 53, 54

Julia Bäckman 49

Sunita Badola 55

Clive Ballard 56

Karina Banasik 57

Nicholas J Bass 6

Aartjan T F Beekman 58

Sintia Belangero 30, 59

Tim B Bigdeli 60

Elisabeth B Binder 50, 61, 62

Ottar Bjerkeset 63, 64

Gyda Bjornsdottir 65

Sigrid Børte 66, 67, 68

Emma Bränn 69

Alice Braun 7

Thorsten Brodersen 70

Tanja M Brückl 71

Søren Brunak 57

Mie T Bruun 72

Margit Burmeister 73

Pichit Buspavanich 74, 75

Jonas Bybjerg-Grauholm 76, 77

Enda M Byrne 78

Jianwen Cai 79

Archie Campbell 80, 81

Megan L Campbell 82

Adrian I Campos 83

Enrique Castelao 84

Jorge Cervilla 85, 86, 87

Boris Chaumette 88

Chia-Yen Chen 89

Hsi-Chung Chen 90, 91

Zhengming Chen 92

Sven Cichon 93, 94, 95, 96

Lucía Colodro-Conde 18, 97

Anne Corbett 56

Elizabeth C Corfield 53, 98

Baptiste Couvy-Duchesne 99

Nick Craddock 100

Udo Dannlowski 52

Gail Davies 101

EJC de Geus 102

Ian J Deary 101

Franziska Degenhardt 94, 103

Abbas Dehghan 104, 105

J Raymond DePaulo 106

Michael Deuschle 5, 107

Maria Didriksen 108

Khoa Manh Dinh 109

Nese Direk 110

Srdjan Djurovic 111, 112

Anna R Docherty 113, 114, 115

Katharina Domschke 116

Joseph Dowsett 108

Ole Kristian Drange 63, 117, 118, 119

Erin C Dunn 10, 120, 121

William Eaton 122

Gudmundur Einarsson 65

Thalia C Eley 8

Samar S M Elsheikh 123

Jan Engelmann 124

Michael E Benros 77, 125, 126

Christian Erikstrup 109

Valentina Escott-Price 100

Chiara Fabbri 8, 127

Yu Fang 73

Sarah Finer 128

Josef Frank 2

Robert C Free 129

Linda Gallo 130

He Gao 131

Michael Gill 132

Maria Gilles 5, 107

Fernando S Goes 106

Scott Douglas Gordon 18, 26

Jakob Grove 43, 47, 48, 133

Daniel F Gudbjartsson 65, 134

Blanca Gutierrez 85, 86, 87

Tim Hahn 52

Lynsey S Hall 1, 100

Thomas F Hansen 57, 135, 136

Magnus Haraldsson 137, 138

Catharina A Hartman 139

Alexandra Havdahl 53, 140

Caroline Hayward 141

Stefanie Heilmann-Heimbach 94

Stefan Herms 93, 94

Ian B Hickie 142

Henrik Hjalgrim 143

Jens Hjerling-Leffler 144

Per Hoffmann 93, 94

Georg Homuth 145

Carsten Horn 146

Jouke-Jan Hottenga 102

David M Hougaard 76, 77

Iiris Hovatta 147

Qin Qin Huang 12

Donald Hucks 39

Floris Huider 102

Karen A Hunt 148

Nicholas S Ialongo 122

Marcus Ising 149

Erkki Isometsä 150

Rick Jansen 58

Yunxuan Jiang 151

Ian Jones 100

Lisa A Jones 152

Lina Jonsson 153

Masahiro Kanai 154, 155, 156

Robert Karlsson 22

Siegfried Kasper 157

Kenneth S Kendler 158

Ronald C Kessler 159

Stefan Kloiber 123, 149, 160, 161

James A Knowles 162

Nastassja Koen 82

Julia Kraft 7

Henry R Kranzler 163, 164

Kristi Krebs 165

Theodora Kunovac Kallak 166

Zoltán Kutalik 167, 168, 169

Elisa Lahtela 170

Marilyn Lake 171

Margit Hørup Larsen 108

Eric J Lenze 172

Melissa Lewins 1

Glyn Lewis 6

Liming Li 173, 174

Bochao Danae Lin 175

Kuang Lin 92

Penelope A Lind 18, 26, 27, 28

Yu-Li Liu 176

Donald J MacIntyre 1

Dean F MacKinnon 106

Brion S Maher 122

Wolfgang Maier 177

Victoria S Marshe 123, 178

Gabriela A Martinez-Levy 179

Koichi Matsuda 180, 181

Hamdi Mbarek 102

Peter McGuffin 8

Sarah E Medland 18, 26, 182, 183

Susanne Meinert 52, 184

Christina Mikkelsen 108, 185

Susan Mikkelsen 109

Yuri Milaneschi 58

Iona Y Millwood 92

Esther Molina 86, 87, 186

Francis M Mondimore 106

Preben Bo Mortensen 24, 42, 43

Benoit H Mulsant 123, 160

Joonas Naamanka 147

Jake M Najman 187

Matthias Nauck 188, 189

Igor Nenadić 190

Kasper R Nielsen 191

Ilja M Nolte 192

Merete Nordentoft 77, 125, 126

Markus M Nöthen 94

Mette Nyegaard 43, 76, 193

Michael C O'Donovan 100

Asmundur Oddsson 65

Adrielle M Oliveira 194

Catherine M Olsen 195, 196

Hogni Oskarsson 197

Sisse Rye Ostrowski 108, 198

Michael J Owen 100

Richard Packer 199

Teemu Palviainen 170

Pedro M Pan 194

Carlos N Pato 200

Michele T Pato 200

Nancy L Pedersen 22

Ole Birger Pedersen 70, 198

Wouter J Peyrot 58

James B Potash 106

Martin Preisig 84

Michael H Preuss 201, 202

Jorge A Quiroz 203

Miguel E Renteria 18, 27, 28

Charles F Reynolds III 204

John P Rice 172

Saori Sakaue 154, 155, 205

Marcos L Santoro 206

Robert A Schoevers 207, 208

Andrew Schork 43, 209

Thomas G Schulze 2, 34, 106, 210, 211, 212

Tabea S Send 107

Jianxin Shi 213

Engilbert Sigurdsson 214

Kritika Singh 39

Grant C B Sinnamon 215

Lea Sirignano 2, 5

Olav B Smeland 119, 216

Daniel J Smith 217

Tamar Sofer 218

Erik Sørensen 108

Sundararajan Srinivasan 219

Hreinn Stefansson 65

Kari Stefansson 65, 138

Peter Straub 39

Mei-Hsin Su 220

André Tadic 124, 221

Henning Teismann 222

Alexander Teumer 223

Anita Thapar 100, 224

Pippa A Thomson 81

Lise Wegner Thørner 108

Apostolia Topaloudi 225

Shih-Jen Tsai 226, 227

Ioanna Tzoulaki 104, 105, 228

George Uhl 229

André G Uitterlinden 230

Henrik Ullum 108, 198, 231

Daniel Umbricht 232

Robert J Ursano 233

Sandra Van der Auwera 223

Albert M van Hemert 234

Abirami Veluchamy 219

Alexander Viktorin 22

Henry Völzke 235

G Bragi Walters 65

Xiaotong Wang 236

Agaz Wani 237

Myrna M Weissman 238

Jürgen Wellmann 222

David C Whiteman 195

Derek Wildman 237

Gonneke Willemsen 102

Alexander T Williams 199

Bendik S Winsvold 67, 68, 239

Stephanie H Witt 2, 5, 240

Ying Xiong 22

Lea Zillich 2

John-Anker Zwart 66, 67, 68

23andMe Research Team 151

China Kadoorie Biobank Collaborative Group 241

Estonian Biobank Research Team 165

Genes & Health Research Team 242

HUNT All-In Psychiatry 243

The BioBank Japan Project 244

VA Million Veteran Program 245

Ole A Andreassen 119, 216, 246

Bernhard T Baune 247, 248, 249

Klaus Berger 222

Dorret I Boomsma 102, 250

Anders D Børglum 43, 47, 48

Gerome Breen 8, 13

Na Cai 251, 252, 253

Hilary Coon 113, 115

William E Copeland 254

Byron Creese 56

Carlos S Cruz-Fuentes 179

Darina Czamara 71

Lea K Davis 39, 255

Eske M Derks 18

Enrico Domenici 256

Paul Elliott 104, 105, 228, 257

Andreas J Forstner 94, 96, 258

Micha Gawlik 259

Joel Gelernter 19, 20, 260

Hans J Grabe 223

Steven P Hamilton 261

Kristian Hveem 67, 262, 263

Catherine John 199, 264

Jaakko Kaprio 170

Tilo Kircher 190

Marie-Odile Krebs 265

Po-Hsiu Kuo 90, 266

Mikael Landén 22, 153

Kelli Lehto 165

Douglas F Levinson 267

Qingqin S Li 268

Klaus Lieb 124

Ruth J F Loos 185, 201, 269, 270, 271

Yi Lu 22

Susanne Lucae 149

Jurjen J Luykx 58, 175, 272

Hermine HM Maes 158, 220, 273

Patrik K Magnusson 22

Hilary C Martin 12

Nicholas G Martin 18, 26

Andrew McQuillin 6

Christel M Middeldorp 78, 274

Lili Milani 165

Ole Mors 43, 275

Daniel J Müller 123, 160, 161, 276

Bertram Müller-Myhsok 50, 277, 278

Yukinori Okada 154, 279, 280

Albertine J Oldehinkel 139

Sara A Paciga 281

Colin NA Palmer 219

Peristera Paschou 225

Brenda WJH Penninx 58

Roy H Perlis 9, 10, 282

Roseann E Peterson 60

Giorgio Pistis 84

Renato Polimanti 19, 32

David J Porteous 81

Danielle Posthuma 283, 284

Jill A Rabinowitz 285

Ted Reichborn-Kjennerud 53

Andreas Reif 44

Frances Rice 100, 224

Roland Ricken 7

Marcella Rietschel 2

Margarita Rivera 86, 87, 286

Christian Rück 49

Giovanni A Salum 287

Catherine Schaefer 288

Srijan Sen 73, 289

Alessandro Serretti 290, 291

Alkistis Skalkidou 166

Jordan W Smoller 9, 292, 293

Dan J Stein 82

Frederike Stein 294

Murray B Stein 295, 296, 297, 298

Patrick F Sullivan 22, 299

Martin Tesli 300

Thorgeir E Thorgeirsson 65

Henning Tiemeier 301, 302

Nicholas J Timpson 25, 303

Monica Uddin 237

Rudolf Uher 304

David A van Heel 148

Karin JH Verweij 305

Robin G Walters 92

Sylvia Wassertheil-Smoller 306

Jens R Wendland 55

Thomas Werge 77, 198, 209, 307, 308

Aeilko H Zwinderman 14

Karoline Kuchenbaecker 6, 92

Naomi R Wray 29, 236, 309

Stephan Ripke 7, 293

Cathryn M Lewis 8, 310

Andrew M McIntosh 1, 81

1, Division of Psychiatry, University of Edinburgh, Edinburgh, UK

2, Department of Genetic Epidemiology in Psychiatry, Central Institute of Mental Health, Medical Faculty Mannheim, Heidelberg University, Mannheim, BW, DE

3, Hector Institute for Artificial Intelligence in Psychiatry, Central Institute of Mental Health, Medical Faculty Mannheim, Heidelberg University, Mannheim, BW, DE

4, Department for Psychiatry and Psychotherapy, Central Institute of Mental Health, Medical Faculty Mannheim, Heidelberg University, Mannheim, BW, DE

5, German Center for Mental Health (DZPG), Partner Site Mannheim - Heidelberg - Ulm, DE

6, Division of Psychiatry, University College London, London, UK

7, Department of Psychiatry and Psychotherapy, Charité – Universitätsmedizin Berlin, Berlin, BE, DE

8, Social, Genetic and Developmental Psychiatry Centre, King's College London, London, UK

9, Department of Psychiatry, Massachusetts General Hospital, Boston, MA, US

10, Department of Psychiatry, Harvard Medical School, Boston, MA, US

11, Department of Clinical and Biomedical Sciences, Faculty of Health and Life Sciences, University of Exeter, Exeter, UK

12, Human Genetics, Wellcome Sanger Institute, Hinxton, UK

13, NIHR Maudsley Biomedical Research Centre, King's College London, London, UK

14, Epidemiologie en Data Science (EDS), Amsterdam UMC, location University of Amsterdam, Amsterdam, NL

15, Institute for Psychopharmacology, Central Institute of Mental Health, Medical Faculty Mannheim, Heidelberg University, Mannheim, BW, DE

16, Department of Psychiatry, College of Health Sciences, University of Alberta, Edmonton, AB, CA

17, Neuroscience and Mental Health Institute, University of Alberta, Edmonton, AB, CA

18, Brain & Mental Health Program, QIMR Berghofer Medical Research Institute, Brisbane, QLD, AU

19, Department of Psychiatry, Yale University School of Medicine, New Haven, CT, US

20, Department of Psychiatry, Veterans Affairs Connecticut Healthcare System, West Haven, CT, US

21, Department of Clinical Medicine, Aarhus University, Aarhus, DK

22, Department of Medical Epidemiology and Biostatistics, Karolinska Institutet, Stockholm, SE

23, Department of Pediatric Neurology, Charité – Universitätsmedizin Berlin, Berlin, BE, DE

24, National Centre for Register-based Research, Aarhus University, Aarhus, DK

25, MRC Integrative Epidemiology Unit, University of Bristol, Bristol, UK

26, Mental Health and Neuroscience, QIMR Berghofer Medical Research Institute, Brisbane, QLD, AU

27, School of Biomedical Sciences, Queensland University of Technology, Brisbane, QLD, AU

28, School of Biomedical Sciences, The University of Queensland, Brisbane, QLD, AU

29, Institute for Molecular Bioscience, University of Queensland, Brisbane, QLD, AU

30, Morphology and Genetics, Universidade Federal de Sao Paulo, Sao Paulo, SP, BR

31, Maurice Wohl Clinical Neuroscience Institute, Department of Basic and Clinical Neuroscience, King's College London, London, UK

32, Veterans Affairs Connecticut Healthcare System, West Haven, CT, US

33, Department of Psychiatry and Psychotherapy, University Hospital, LMU Munich, Munich, BY, DE

34, Institute of Psychiatric Phenomics and Genomics, University Hospital, LMU Munich, Munich, BY, DE

35, Department of Psychiatry and Psychotherapy, University Hospital Bonn, Medical Faculty, University of Bonn, Bonn, DE

36, Institute of Human Genetics, University Hospital Bonn, Medical Faculty, University of Bonn, Bonn, DE

37, German Center for Mental Health (DZPG), Partner Site Munich - Augsburg, DE

38, Old Age Psychiatry, King's College London, London, UK

39, Department of Medicine, Division of Genetic Medicine, Vanderbilt University Medical Center, Nashville, TN, US

40, Department of Psychiatry and Psychotherapy, Charité – Universitätsmedizin Berlin, Campus Charité Mitte (CCM), Berlin, BE, DE

41, Center for Psychiatry, Psychotherapy and Psychosomatic Medicine, Fliedner Klinik Berlin, Berlin, BE, DE

42, Centre for Integrated Register-based Research, Aarhus University, Aarhus, DK

43, iPSYCH, The Lundbeck Foundation Initiative for Integrative Psychiatric Research, Aarhus, DK

44, Department of Psychiatry, Psychosomatic Medicine and Psychotherapy, Goethe University Frankfurt - University Hospital, Frankfurt am Main, DE

45, Department of Epidemiology, Columbia University Mailman School of Public Health, New York, NY, US

46, Discipline of Psychiatry, University of Adelaide, Adelaide, SA, AU

47, Department of Biomedicine and Centre for Integrative Sequencing, iSEQ, Aarhus University, Aarhus, DK

48, Center for Genomics and Personalized Medicine, Aarhus University, Aarhus, DK

49, Department of Clinical Neuroscience, Karolinska Institutet, Stockholm, SE

50, Department of Translational Research in Psychiatry, Max Planck Institute of Psychiatry, Munich, BY, DE

51, Department of Neurology, Klinikum rechts der Isar, Technical University of Munich, Munich, BY, DE

52, Institute for Translational Psychiatry, University of Münster, Münster, NRW, DE

53, PsychGen Centre for Genetic Epidemiology and Mental Health, Norwegian Institute of Public Health, Oslo, OSL, NO

54, PROMENTA Research Center, Department of Psychology, University of Oslo, Oslo, OSL, NO

55, Research and Development, Takeda Pharmaceutical Company Limited, Cambridge, MA, US

56, Faculty of Health and Life Sciences, University of Exeter, Exeter, UK

57, Novo Nordisk Foundation Center for Protein Research, Faculty of Health and Medical Sciences, University of Copenhagen, Copenhagen, CPH, DK

58, Department of Psychiatry, Amsterdam Public Health and Amsterdam Neuroscience, Amsterdam UMC, Vrije Universiteit Amsterdam, Amsterdam, NL

59, Laboratory of Integrative Neuroscience, Universidade Federal de Sao Paulo, Sao Paulo, SP, BR

60, Department of Psychiatry and Behavioral Sciences, Institute for Genomics in Health, State University of New York Downstate Health Sciences University, Brooklyn, NY, US

61, Department of Psychiatry and Behavioral Sciences, Emory University School of Medicine, Atlanta, GA, US

62, Department Genes and Environment, Max Planck Institute of Psychiatry, Munich, BY, DE

63, Department of Mental Health, Faculty of Medicine and Health Sciences, Norwegian University of Science and Technology (NTNU), Trondheim, TRD, NO

64, Faculty of Nursing and Health Sciences, NORD University, Levanger, NO

65, deCODE Genetics / Amgen, Reykjavik, IS

66, Institute of Clinical Medicine, Faculty of Medicine, University of Oslo, Oslo, OSL, NO

67, HUNT Center for Molecular and Clinical Epidemiology, Department of Public Health and Nursing, Faculty of Medicine and Health Sciences, Norwegian University of Science and Technology, Trondheim, TRD, NO

68, Department of Research and Innovation, Division of Clinical Neuroscience, Oslo University Hospital, Oslo, OSL, NO

69, Institute of Environmental Medicine, Unit of Integrative Epidemiology, Karolinska Institutet, Stockholm, SE

70, Department of Clinical Immunology, Zealand University Hospital, Køge, DK

71, Department Genes and Environment, Max Planck Institute of Psychiatry, Munich, BY, DE

72, Department of Clinical Immunology, Odense University Hospital, Odense, DK

73, Michigan Neuroscience Institute, University of Michigan, Ann Arbor, MI, US

74, Department of Psychiatry and Psychotherapy, Gender Research in Medicine, Institute of Sexology and Sexual Medicine, Charité – Universitätsmedizin Berlin, Berlin, BE, DE

75, Department of Psychiatry, Psychotherapy and Psychosomatics, Brandenburg Medical School Theodor Fontane, Neuruppin, BB, DE

76, Center for Neonatal Screening, Department for Congenital Disorders, Statens Serum Institut, Copenhagen, CPH, DK

77, iPSYCH, The Lundbeck Foundation Initiative for Integrative Psychiatric Research, Copenhagen, CPH, DK

78, Child Health Research Centre, University of Queensland, Brisbane, QLD, AU

79, Department of Biostatistics , University of North Carolina at Chapel Hill  , Chapel Hill, NC, US

80, Centre for Medical Informatics, Usher Institute, University of Edinburgh, Edinburgh, UK

81, Centre for Genomic & Experimental Medicine, Institute for Genetics and Cancer, University of Edinburgh, Edinburgh, UK

82, SAMRC Unit on Risk & Resilience in Mental Disorders, Department of Psychiatry and Neuroscience Institute, University of Cape Town, Cape Town, SA

83, Statistical genetics, Institute for Molecular Bioscience, The University of Queensland, Brisbane, QLD, AU

84, Department of Psychiatry, Lausanne University Hospital and University of Lausanne, Prilly, VD, CH

85, Department of Psychiatry, Faculty of Medicine, University of Granada, Granada, ES

86, Instituto de Investigación Biosanitaria, Ibs Granada, Granada, ES

87, Institute of Neurosciences ´Federico Olóriz´, Biomedical Research Centre (CIBM), University of Granada, Granada, ES

88, Université de Paris Cité, INSERM U1266, Institute of Psychiatry and Neuroscience of Paris, GHU Paris Psychiatry and Neuroscience, Paris, FR

89, Biogen, Cambridge, MA, US

90, Department of Psychiatry, National Taiwan University Hospital,, TW

91, School of Medicine, National Taiwan University College of Medicine, Taipei, TW

92, Nuffield Department of Population Health, University of Oxford, Oxford, UK

93, Human Genomics Research Group, Department of Biomedicine, University of Basel, Basel, CH

94, Institute of Human Genetics, University of Bonn, School of Medicine & University Hospital Bonn, Bonn, DE

95, Institute of Medical Genetics and Pathology, University Hospital Basel, University of Basel, Basel, CH

96, Institute of Neuroscience and Medicine (INM-1), Research Center Juelich, Juelich, DE

97, School of Psychology, University of Queensland, Brisbane, QLD, AU

98, Nic Waals Institute, Lovisenberg Diakonale Hospital, Oslo, OSL, NO

99, Centre for Advanced Imaging, University of Queensland, Saint Lucia, QLD, AU

100, Centre for Neuropsychiatric Genetics and Genomics, Cardiff University, Cardiff, UK

101, The Lothian Birth Cohorts, University of Edinburgh, Edinburgh, UK

102, Department of Biological Psychology & Amsterdam Public Health Research Institute, Vrije Universiteit Amsterdam, Amsterdam, NL

103, Department of Child and Adolescent Psychiatry, Psychosomatics and Psychotherapy, University Hospital Essen, University of Duisburg-Essen, Duisburg, DE

104, MRC Centre for Environment and Health, School of Public Health, Imperial College London, London, UK

105, Imperial College Dementia Research Institute, Imperial College London, London, UK

106, Department of Psychiatry and Behavioral Sciences, Johns Hopkins University School of Medicine, Baltimore, MD, US

107, Department of Psychiatry and Psychotherapy, Research Group Stress Related Disorders, Central Institute of Mental Health, Medical Faculty Mannheim, Heidelberg University, Mannheim, BW, DE

108, Department of Clinical Immunology, Copenhagen University Hospital, Rigshospitalet, Copenhagen, CPH, DK

109, Department of Clinical Immunology, Aarhus University Hospital, Aarhus, DK

110, Department of Psychiatry, Istanbul University, Istanbul, TR

111, Department of Medical Genetics, Oslo University Hospital, Oslo, OSL, NO

112, NORMENT, Department of Clinical Science, University of Bergen, Bergen, NO

113, Psychiatry, University of Utah School of Medicine, Salt Lake City, UT, US

114, Center for Genomic Medicine, Salt Lake City, UT, US

115, Huntsman Mental Health Institute, Salt Lake City, UT, US

116, Department of Psychiatry and Psychotherapy, Medical Center, University of Freiburg, Faculty of Medicine, University of Freiburg, Freiburg, DE

117, Division of Mental Health Care, St. Olavs Hospital, Trondheim University Hospital, Trondheim, TRD, NO

118, Department of Psychiatry, Sørlandet Hospital, Kristiansand, AG, NO

119, NORMENT, Institute of Clinical Medicine, University of Oslo, Oslo, OSL, NO

120, Center for Genomic Medicine, Massachusetts General Hospital, Boston, MA, US

121, Department of Sociology, Purdue University, West Lafayette, IN, US

122, Department of Mental Health, Johns Hopkins, Baltimore, MD, US

123, Centre for Addiction and Mental Health, Toronto, ON, CA

124, Department of Psychiatry and Psychotherapy, University Medical Center of the Johannes Gutenberg University Mainz, Mainz, DE

125, Mental Health Center Copenhagen, Mental Health Services Capital Region of Denmark, Copenhagen, CPH, DK

126, Faculty of Health Science, Department of Clinical Medicine, University of Copenhagen, Copenhagen, CPH, DK

127, Department of Biomedical and Neuromotor Sciences, University of Bologna, Bologna, IT

128, Wolfson Institute of Population Health, Queen Mary University of London, London, UK

129, School of Computing and Mathematical Sciences, University of Leicester, Leicester, UK

130, Department of Psychology, San Diego San Diego State University, San Diego, CA, US

131, Department of Epidemiology and Biostatistics, Imperial College London, London, UK

132, Discipline of Psychiatry, School of Medicine, Trinity College Dublin, Dublin, IE

133, Bioinformatics Research Centre, Aarhus University, Aarhus, DK

134, School of Engineering, University of Iceland, Reykjavik, IS

135, Danish Headache Centre, Department of Neurology, Rigshospitalet, Glostrup, DK

136, Neurogenomics Group, Translational Research Centre, Rigshospitalet Copenhagen University Hospital, Glostrup, DK

137, Landspitali-University Hospital, Reykjavik, IS

138, Faculty of Medicine, University of Iceland, Reykjavik, IS

139, Department of Psychiatry, University of Groningen, University Medical Center Groningen, Groningen, NL

140, Nic Waals Institute, Lovisenberg Diaconal Hospital, Oslo, OSL, NO

141, MRC Human Genetics Unit, Institute for Genetics and Cancer, University of Edinburgh, Edinburgh, UK

142, Brain and Mind Centre, University of Sydney, Sydney, NSW, AU

143, Department of Epidemiology Research, Statens Serum Institut, Copenhagen, CPH, DK

144, Department of Medical Biochemistry and Biophysics, Karolinska Institutet, Stockholm, SE

145, Interfaculty Institute for Genetics and Functional Genomics, Department of Functional Genomics, University Medicine Greifswald, Greifswald, MV, DE

146, Roche Pharmaceutical Research and Early Development, Pharmaceutical Sciences, Roche Innovation Center Basel, F. Hoffmann-La Roche Ltd, Basel, CH

147, SleepWell Research Program and Department of Psychology and Logopedics, University of Helsinki, Helsinki, FI

148, Blizard Institute, Barts and the London School of Medicine and Dentistry, Queen Mary University of London, London, UK

149, Max Planck Institute of Psychiatry, Munich, BY, DE

150, Department of Psychiatry, University of Helsinki, Helsinki, FI

151, 23andMe Research Team, 23andMe, Inc., Sunnyvale, CA, US

152, Department of Psychological Medicine, University of Worcester, Worcester, UK

153, Institution of Neuroscience and Physiology, University of Gothenburg, Gothenburg, SE

154, Department of Statistical Genetics, Osaka University Graduate School of Medicine, Suita, JP

155, Program in Medical and Population Genetics, Broad Institute of Harvard and MIT, Cambridge, MA, US

156, Center for Computational and Integrative Biology, Massachusetts General Hospital, Boston, MA, US

157, Center for Brain Research, Department of Molecular Neuroscience, Medical University of Vienna, Vienna, AT

158, Department of Psychiatry, Virginia Commonwealth University, Richmond, VA, US

159, Department of Health Care Policy, Harvard Medical School, Boston, MA, US

160, Department of Psychiatry, University of Toronto, Toronto, ON, CA

161, Department of Pharmacology & Toxicology, University of Toronto, Toronto, ON, CA

162, Department of Genetics, Rutgers University, Piscataway, NJ, US

163, Department of Psychiatry, Perelman School of Medicine, University of Pennsylvania, Philadelphia, PA, US

164, Mental Illness Research, Education and Clinical Center, Crescenz VA Medical Center, Philadelphia, PA, US

165, Estonian Genome Centre, Institute of Genomics, University of Tartu, Tartu, EE

166, Department of Women's and Children's Health, Uppsala University, Uppsala, SE

167, Department of Epidemiology and Health Systems, Center for Primary Care and Public Health, Lausanne, VD, CH

168, Department of Computational Biology, University of Lausanne, Lausanne, VD, CH

169, Swiss Institute of Bioinformatics, Lausanne, VD, CH

170, Institute for Molecular Medicine Finland - FIMM, University of Helsinki, Helsinki, FI

171, SAMRC Unit on Risk & Resilience in Mental Disorders, Department of Psychiatry and Neuroscience Institute, University of Cape Town, Cape Town, SA

172, Department of Psychiatry, Washington University School of Medicine in St. Louis, St. Louis, MO, US

173, Department of Epidemiology and Biostatistics, School of Public Health, Peking University, Beijing, CN

174, Peking University Center for Public Health and Epidemic Preparedness & Response, Peking University, Beijing, CN

175, Department of Psychiatry and Neuropsychology, School for Mental Health and Neuroscience, Maastricht University Medical Centre, Maastricht, NL

176, Center for Neuropsychiatric Research, National Health Research Institutes,, TW

177, Department of Psychiatry and Psychotherapy, University of Bonn, Bonn, DE

178, Center for Translational and Computational Neuroimmunology, Columbia University Medical Center, New York, NY, US

179, Psychiatric Genetics Department, Instituto Nacional de Psiquiatría Ramón de la Fuente Muñiz (INPRFM), Mexico City, CDMX, MX

180, Laboratory of Genome Technology, Human Genome Center, Institute of Medical Science, The University of Tokyo, Tokyo, JP

181, Laboratory of Clinical Genome Sequencing, Department of Computational Biology and Medical Sciences, Graduate School of Frontier Sciences, The University of Tokyo, Tokyo, JP

182, School of Psychology, The University of Queensland, Brisbane, QLD, AU

183, School of Psychology and Counselling, Queensland University of Technology, Brisbane, QLD, AU

184, Institute for Translational Neuroscience, University of Münster, Münster, NRW, DE

185, Novo Nordisk Foundation Center for Basic Metabolic Research, Faculty of Health and Medical Sciences, University of Copenhagen, Copenhagen, CPH, DK

186, Department of Nursing, Faculty of Health Sciences, University of Granada, Granada, ES

187, School of Public Health, University of Queensland, Brisbane, QLD, AU

188, Institute of Clinical Chemistry and Laboratory Medicine, University Medicine Greifswald, Greifswald, MV, DE

189, DZHK (German Centre for Cardiovascular Research), Partner Site Greifswald, Greifswald, MV, DE

190, Department of Psychiatry, University of Marburg, Marburg, HE, DE

191, Department of Clinical Immunology, Aalborg University Hospital, Aalborg, DK

192, Department of Epidemiology, University of Groningen, University Medical Center Groningen, Groningen, NL

193, Department of Health, Science and Technology, Aalborg University, Aalborg, DK

194, Department of Psychiatry, Universidade Federal de Sao Paulo, Sao Paulo, SP, BR

195, Population Health, QIMR Berghofer Medical Research Institute, Brisbane, QLD, AU

196, The Fraser Institute, Faculty of Medicine, University of Queensland, Brisbane, QLD, AU

197, Directorate of Health, Iceland, Reykjavik, IS

198, Department of Clinical Medicine, University of Copenhagen, Copenhagen, CPH, DK

199, Department of Population Health Sciences, University of Leicester, Leicester, UK

200, Department of Psychiatry, Rutgers University, Piscataway, NJ, US

201, Charles Bronfman Institute for Personalized Medicine, Icahn School of Medicine at Mount Sinai, New York, NY, US

202, Department of Environmental Medicine and Public Health, Icahn School of Medicine at Mount Sinai, New York, NY, US

203, Translational Medicine, Roche, New York, NY, US

204, Psychiatry, University of Pittsburgh Medical Centre, Pittsburgh, PA, US

205, Divisions of Genetics and Rheumatology, Department of Medicine, Brigham and Women’s Hospital, Harvard Medical School, Boston, MA, US

206, Department of Biochemistry, Universidade Federal de Sao Paulo, Sao Paulo, SP, BR

207, Department of Psychiatry, University Medical Center Groningen, Groningen, NL

208, Research School of Behavioural and Cognitive Neurosciences (BCN), University of Groningen, Groningen, NL

209, Institute of Biological Psychiatry, Mental Health Center Sct. Hans, Mental Health Services Capital Region of Denmark, Copenhagen, CPH, DK

210, Department of Psychiatry and Psychotherapy, University Medical Center Göttingen, Goettingen, NI, DE

211, Human Genetics Branch, NIMH Division of Intramural Research Programs, Bethesda, MD, US

212, Department of Psychiatry and Behavioral Sciences, SUNY Upstate Medical University, Syracuse, NY, USA

213, Division of Cancer Epidemiology and Genetics, National Cancer Institute, Bethesda, MD, US

214, Faculty of Medicine, Department of Psychiatry, University of Iceland, Reykjavik, IS

215, School of Medicine and Dentistry, James Cook University, Townsville, QLD, AU

216, Division of Mental Health and Addiction, Oslo University Hospital, Oslo, OSL, NO

217, Center for Clinical Brain Sciences, University of Edinburgh, Edinburgh, UK

218, Beth Israel Deaconess Medica, Harvard Medical School, Boston, MA, US

219, Division of Population Health and Genomics, Ninewells Hospital and School of Medicine, University of Dundee, Dundee, UK

220, Virginia Institute for Psychiatric and Behavioral Genetics, Virginia Commonwealth University, Richmond, VA, US

221, Department of Psychiatry, Psychotherapy and Psychosomatics, Dr. Fontheim Mentale Gesundheit, Liebenburg, DE

222, Institute of Epidemiology and Social Medicine, University of Münster, Münster, NRW, DE

223, Department of Psychiatry and Psychotherapy, University Medicine Greifswald, Greifswald, MV, DE

224, Wolfson Centre for Young People's Mental Health, Division of Psychological Medicine and Clinical Neurosciences, Cardiff University, Cardiff, UK

225, Department of Biological Sciences, Purdue University, West Lafayette, IN, US

226, Institute of Brain Science & Division of Psychiatry, National Yang-Ming University,, TW

227, Department of Psychiatry, Taipei Veterans General Hospital, Taipei, ROC, TW

228, Imperial College BHF Centre for Research Excellence, Imperial College London, London, UK

229, University of Maryland School of Medicine and VA Maryland Healthcare System, Baltimore, MD, US

230, Department of Internal Medicine, Erasmus University Medical Center Rotterdam, Rotterdam, NL

231, Management Section, Statens Serum Institut, Copenhagen, CPH, DK

232, Xperimed LLC, Basel, CH

233, Department of Psychiatry, Uniformed Services University of the Health Sciences, Bethesda, MD, US

234, Department of Psychiatry, Leiden University Medical Center, Leiden, NL

235, Institute for Community Medicine, University Medicine Greifswald, Greifswald, MV, DE

236, Department of Psychiatry, University of Oxford, Oxford, UK

237, Genomics Program, University of South Florida College of Public Health, Tampa, FL, US

238, Columbia University Vagelos College of Physicians and Surgeons, New York, NY, US

239, Department of Neurology, Oslo University Hospital, Oslo, OSL, NO

240, Center for Innovative Psychiatric and Psychotherapeutic Research, Central Institute of Mental Health, Medical Faculty Mannheim, Heidelberg University, Mannheim, BW, DE

241, China Kadoorie Biobank Collaborative Group

242, Genes & Health Research Team

243, HUNT All-In Psychiatry

244, The BioBank Japan Project

245, VA Million Veteran Program

246, KG Jebsen Centre for Neurodevelopmental Research, University of Oslo, Oslo, OSL, NO

247, Department of Psychiatry, University of Münster, Münster, NRW, DE

248, Department of Psychiatry, University of Melbourne, Melbourne, VIC, AU

249, Florey Institute of Neuroscience and Mental Health, University of Melbourne, Melbourne, VIC, AU

250, Department of Complex Trait Genetics, CNCR, Vrije Universiteit Amsterdam, Amsterdam, NL

251, Helmholtz Pioneer Campus, Helmholtz Zentrum München, Neuherberg, DE

252, Computational Health Centre, Helmholtz Zentrum München, Neuherberg, DE

253, School of Medicine, Technical University of Munich, Munich, BY, DE

254, Department of Psychiatry, University of Vermont, Burlington, VT, US

255, Department of Medicine, Icahn School of Medicine at Mount Sinai, New York, NY, US

256, Department of Cellular, Computational and Integrative Biology, Università degli Studi di Trento, Trento, IT

257, Imperial College Biomedical Research Centre, Imperial College London, London, UK

258, Center for Human Genetics, University of Marburg, Marburg, HE, DE

259, Department of Psychiatry, Psychosomatics and Psychotherapy, Julius-Maximilians-Universität Würzburg, Würzburg, DE

260, Department of Genetics, Department of Neuroscience, Yale University School of Medicine, New Haven, CT, US

261, Psychiatry, Kaiser Permanente Northern California, San Francisco, CA, US

262, HUNT Research Center, Department of Public Health and Nursing, Faculty of Medicine and Health Sciences, Norwegian University of Science and Technology (NTNU), Trondheim, NO

263, Department of Research, Innovation and Education, St. Olavs Hospital, Trondheim University Hospital, Trondheim, TRD, NO

264, University Hospitals of Leicester NHS Trust, Leicester, UK

265, Pathophysiology of Psychiatric Diseases, INSERM, Univ Paris Cité, GHU Paris, Paris, FR

266, Institute of Epidemiology and Preventive Medicine & Department of Public Health, National Taiwan University,, TW

267, Department of Psychiatry & Behavioral Sciences, Stanford University, Stanford, CA, US

268, Neuroscience Therapeutic Area, Janssen Research and Development, LLC, Titusville, NJ, US

269, Mindich Child Health and Development Institute, Icahn School of Medicine at Mount Sinai, New York, NY, US

270, Department of Environmental Medicine and Public Health, Icahn School of Medicine at Mount Sinai, New York, NY, US

271, MRC Metabolic Diseases Unit, University of Cambridge Metabolic Research Laboratories, Wellcome-MRC Institute of Metabolic Science, Addenbrooke's Hospital, Cambridge, UK

272, Bipolar Disorders Outpatient Clinic, GGZ InGeest, Amsterdam, NL

273, Department of Human and Molecular Genetics, Virginia Commonwealth University, Richmond, VA, US

274, Child and Youth Mental Health Service, Children's Health Queensland Hospital and Health Service, Brisbane, QLD, AU

275, Psychosis Research Unit, Aarhus University Hospital-Psychiatry, Aarhus, DK

276, Department of Psychiatry, Psychosomatics and Psychotherapy, University Hospital of Würzburg, Würzburg, DE

277, Munich Cluster for Systems Neurology (SyNergy), Munich, BY, DE

278, University of Liverpool, Liverpool, UK

279, Department of Genome Informatics, Graduate School of Medicine, The University of Tokyo, Tokyo, JP

280, Laboratory for Systems Genetics, RIKEN Center for Integrative Medical Sciences, Yokohama, JP

281, Human Genetics and Computational Biomedicine, Pfizer Global Research and Development, Groton, CT, US

282, Centre for Quantitative Health, Massachusetts General Hospital, Boston, MA, US

283, Child and Adolescent Psychiatry, Amsterdam UMC, Vrije Universiteit Amsterdam, Amsterdam, NL

284, Complex Trait Genetics, Vrije Universiteit Amsterdam, Amsterdam, NL

285, Department of Mental Health, Johns Hopkins University, Baltimore, MD, US

286, Department of Biochemistry and Molecular Biology II, Faculty of Pharmacy, University of Granada, Granada, ES

287, Psychiatry, Universidade Federal do Rio Grande do Sul, Porto Alegre, BR

288, Division of Research, Kaiser Permanente Northern California, Oakland, CA, US

289, Eisenberg Family Depression Center, University of Michigan, Ann Arbor, MI, US

290, Department of Medicine and Surgery, Kore University of Enna, Enna, IT

291, Psychiatry, Oasi Research Institute-IRCCS, Troina, IT

292, Psychiatric and Neurodevelopmental Genetics Unit, Massachusetts General Hospital, Boston, MA, US

293, Stanley Center for Psychiatric Research, Broad Institute of MIT and Harvard, Cambridge, MA, US

294, Department of Psychiatry and Psychotherapy, University of Marburg, Marburg, HE, DE

295, Psychiatry Service, Veterans Affairs San Diego Healthcare System, San Diego, CA, US

296, School of Public Health, University of California, San Diego, La Jolla, CA, US

297, Department of Psychiatry, University of California, San Diego, La Jolla, CA, US

298, Psychiatry, Veterans Affairs San Diego Healthcare System, San Diego, CA, US

299, Departments of Genetics and Psychiatry, University of North Carolina at Chapel Hill, Chapel Hill, NC, US

300, Department of Mental Health and Suicide, Norwegian Institute of Public Health, Oslo, OSL, NO

301, Child and Adolescent Psychiatry, Erasmus University Medical Center Rotterdam, Rotterdam, NL

302, Social and Behavioral Science, Harvard T.H. Chan School of Public Health, Boston, MA, US

303, Population Health Sciences, Bristol Medical School, University of Bristol, Bristol, UK

304, Psychiatry, Dalhousie University, Halifax, NS, CA

305, Psychiatry, Amsterdam UMC, location University of Amsterdam, Amsterdam, NL

306, Epidemiology and Population Health, Albert Einstein College of Medicine, Bronx, NY, US

307, Institute of Biological Psychiatry, Mental Health Center Sct. Hans, Copenhagen University Hospital, Mental Health Services, Copenhagen, CPH, DK

308, GLOBE Institute, Lundbeck Foundation Centre for Geogenetics, University of Copenhagen, Copenhagen, CPH, DK

309, Queensland Brain Institute, University of Queensland, Brisbane, QLD, AU

310, Department of Medical & Molecular Genetics, King's College London, London, UK
